# IgA deficiency destabilizes immunological homeostasis towards intestinal microbiota and increases the risk of systemic immune dysregulation

**DOI:** 10.1101/2021.08.20.21261620

**Authors:** Peyton E. Conrey, Lidiya Denu, Kaitlin C. O’Boyle, Jamal Green, Jeffrey Maslanka, Jean-Bernard Lubin, Tereza Duranova, Brittany L. Haltzman, Lauren Gianchetti, Derek Oldridge, Laura A Vella, David Allman, Jonathan Spergel, Ceylan Tanes, Kyle Bittinger, Sarah E. Henrickson, Michael A. Silverman

## Abstract

Mammals produce large quantities of mucosal and systemic antibodies that maintain the intestinal barrier, shape the intestinal microbiome and promote lifelong mutualism with commensal microbes. Here, we developed an integrated host-commensal approach combining microbial flow cytometry and 16s rRNA gene sequencing to define the core microbes that induce mucosal and systemic antibodies in pediatric selective Immunoglobulin A (IgA) deficient and household control siblings with CyTOF analysis to determine the impacts of IgA deficiency on host cellular immune phenotype. In healthy controls, mucosal secretory IgA and IgM antibodies coat an overlapping subset of microbes, predominantly Firmicutes and Proteobacteria. Serum IgG antibodies target a similar consortium of fecal microbes, revealing connections between mucosal and systemic antibody networks. Unexpectedly, IgM provides limited compensation for IgA in children lacking intestinal IgA. Furthermore, we find broad systemic immune dysregulation in a subset of children and mice lacking IgA, including enhanced IgG targeting of fecal microbiota, elevated levels of inflammatory and allergic cytokines and alterations in T cell activation state. Thus, IgA tunes systemic interactions between the host and commensal microbiota. Understanding how IgA tunes baseline immune tone has implications for predicting and preventing autoimmune, inflammatory and allergic diseases broadly, as well as providing improved prognostic guidance to patients with IgA deficiency.

**One Sentence Summary:** IgA deficiency impairs immune homeostasis toward microbiota in children, increasing the risk of immune dysregulation.

## Introduction

Mammals evolved an efficient, layered immune system to prevent infection while maintaining homeostasis with commensal microbes(Herzenberg and Herzenberg, 1989). To that end, mammals invest heavily in secretory mucosal antibodies, producing abundant quantities of secretory dimeric IgA (sIgA)(Bakema and van Egmond, 2011) which enter the intestinal lumen and bind commensal microbes(Catanzaro et al., 2019; Fadlallah et al., 2018; Magri et al., 2017). Historically, the primary role of mucosal IgA was thought to be strict containment of pathogenic and commensal microbes within non-sterile sites, such as the intestine(Mantis et al., 2011; Stokes et al., 1975). However, recent studies provide multiple lines of evidence which challenge the concept of strict immune exclusion, instead supporting a model of “homeostatic immunity” in which the immune system maintains an active, yet non-inflammatory, stance towards commensal microbes at non-sterile barrier sites(Ansaldo et al., 2021; Belkaid and Harrison, 2017). First, live commensal microbes translocate at a low level to the systemic circulation(Lockhart et al., 2008), and colonize lymphoid tissues, without disrupting homeostasis(Obata et al., 2010). Second, microbial products are rapidly and widely distributed throughout mammalian tissues (Uchimura et al., 2018). Third, circulating IgG antibodies and CD4 T cells that recognize commensal microbes are readily found in the systemic circulation of mice and humans(Hegazy et al., 2017; Koch et al., 2016; Zeng et al., 2016). Taken together these findings suggest an intimate and important awareness of commensal microbes and their antigens by systemic immune components, yet the degree to which IgA, the main secretory antibody, impacts the mucosal and systemic dialogue between host and commensal microbes remains unclear.

A long-standing paradox has been the apparently central role of sIgA in mucosal biology alongside the rather mild phenotype in most IgA deficient humans and mice. Selective IgA deficiency (SIgAD) is the most common primary immune deficiency affecting ∼1 in 600 Caucasian individuals(Jorgensen et al., 2013; Yel, 2010). In this disorder, patients over 4 years of age have absent serum IgA and normal serum levels of IgG and IgM, without other known components of immune deficiency. Patients have variable clinical presentations, with a subset of patients developing recurrent infections, severe atopy or autoimmune disease, while others remain apparently asymptomatic(Jorgensen et al., 2013; Lougaris et al., 2019; Yel, 2010). Various compensatory mechanisms for IgA deficiency, including increased secretory and circulating IgM, have been proposed(Catanzaro et al., 2019; Fadlallah et al., 2018; Macpherson et al., 2018). However, it remains unclear how sIgA and sIgM work together in the mucosal system, and on a larger scale, whether the systemic and mucosal anti-commensal networks are redundant or possess unique features. Further, we lack an understanding of why IgA deficiency predisposes some individuals to develop infection or systemic immune dysregulation.

Advances in fluorescence activated cell sorting and microbiome sequencing technologies have helped define some of the microbial targets of secretory IgA, secretory IgM and circulating IgG in humans(Catanzaro et al., 2019; Fadlallah et al., 2018; Fadlallah et al., 2019; Janzon et al., 2019; Magri et al., 2017), but several key questions remain, and we endeavored to learn from pediatric SIgAD to better understand the basic immune mechanisms of IgA function and interplay with the gut microbiota and cellular immunity. First, what are the unique and potentially redundant roles of secretory IgA and IgM? Second, what is the relationship between anti-commensal responses in the mucosal (i.e., sIgA and sIgM) and systemic compartments (i.e., IgG)? Third, what are the impacts of IgA deficiency on systemic immune homeostasis? Should there be alterations in the community of commensal microbes or increased access of these microbes to systemic immune sites, there is the potential for the development of immune dysregulation. To address these questions, we developed an integrated host-commensal approach combining microbial flow cytometry and 16s rRNA gene sequencing (mFLOW-Seq) with CyTOF analysis of peripheral blood mononuclear cells (PBMCs) to simultaneously define the microbial targets of intestinal sIgA and sIgM and circulating IgM and IgG in a cohort of children with SIgAD and their IgA sufficient siblings to determine the impacts of IgA deficiency on host immune state. We show that mucosal and systemic antibody networks cooperate to maintain homeostasis by targeting a common subset of commensal microbes, and that IgA provides non-redundant functions to restrain systemic responses to these commensals in both mice and humans.

## Results

### Secretory IgA and IgM coat an overlapping subset of commensal microbes enriched in Firmicutes

To investigate mucosal and systemic antibody responses to intestinal microbiota in children, we collected fecal and blood samples from sets of pediatric siblings from 15 families that included at least one sibling diagnosed with SIgAD. Since maternal, genetic and environmental factors shape commensal microbiota and immune system development(Ansaldo et al., 2021; Atyeo and Alter, 2021; Brodin and Davis, 2017), we employed a sibling-controlled study design to minimize differences due to these important variables. Age and sex were evenly distributed in SIgAD and sibling controls (**Table S1**). From these 15 families, all 15 probands with the diagnosis of SIgAD had serum IgA levels <7 mg/dL, and 13/17 siblings had normal or slightly low, age-adjusted IgA. Unexpectedly 4 control siblings had undetectable serum IgA, which will be discussed below.

To determine the proportion of gut microbes bound by antibodies, we performed microbial flow cytometry (mFLOW)(Koch et al., 2016). In mFLOW, isolated fecal microbes were stained with the nucleic acid dye SYTO BC and fluorophore-conjugated polyclonal secondary antibodies specific for the Fc portion of human IgA, IgM and IgG. Samples were analyzed by standard flow cytometry using low side scatter (SSC) and forward scatter (FSC) parameters (Fig 1A). In this cohort, IgA sufficient siblings’ secretory IgA (sIgA) and IgM (sIgM) bound to 19.9% and 25.8% of the fecal microbiota, respectively, while IgG did not coat fecal microbes, as expected (Fig 1A-B). 10.7% of fecal microbes were dually coated by sIgA and sIgM, while 8.9% and 11.1% of fecal microbes were uniquely bound by either IgA or IgM, respectively (**Fig S1A**). This pattern of binding of secretory IgM and IgA to commensals suggests two general models: 1) a relatively small subset of microbial species are reliably bound by sIgA **or** sIgM, or 2) many commensal microbes are bound to some degree by sIgA **and** sIgM. In the first model, specific microbes are discretely categorized as inducing sIgM or sIgA, while the second model suggests that both secretory antibodies target most commensal microbes to some degree.

**Figure 1.**
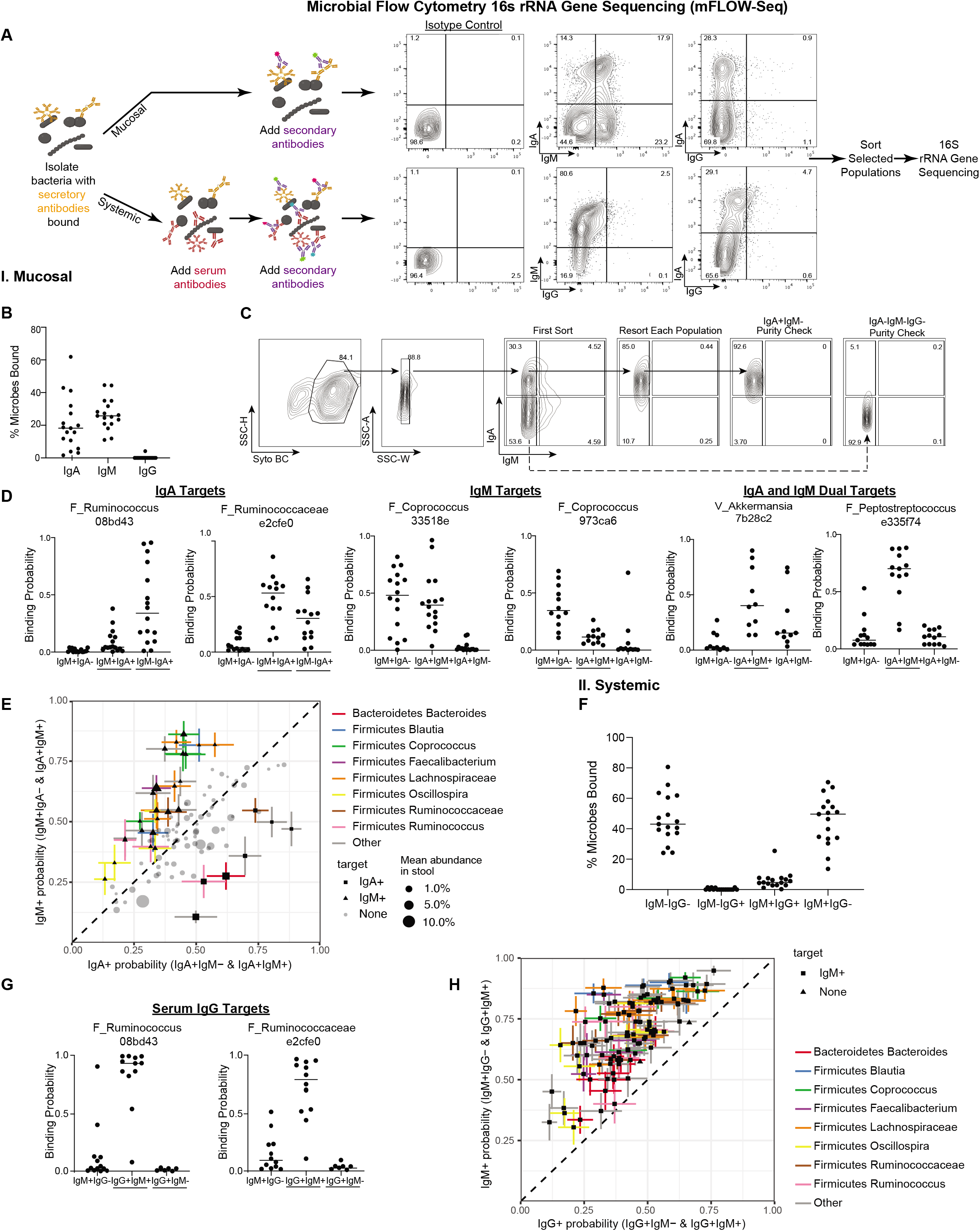
Secretory IgA and IgM and systemic IgG coat diverse fecal microbes in healthy children. **A-B.** Microbial flow cytometry and 16s rRNA gene sequencing (mFLOW-Seq) schematic and gating scheme. Fecal microbes were incubated with or without autologous serum (normalized to 10 ug/mL of IgG) stained with the nucleic acid dye Syto BC and either secondary fluorophore conjugated anti-IgA, anti-IgM or anti-IgG antibodies or isotype controls and analyzed by flow cytometry or sorted into selected population. 16s rRNA gene sequencing and microbiome analysis is performed on sorted populations. **C.** Microbial flow cytometric cell sorting schematic. 100,000 cells from each population were sorted twice and then checked for purity of the desired populations. This example depicts sorting for IgA^-^IgM^-^ and IgA^+^IgM^-^ populations from a single subject. **D.** Binding probabilities of representative microbes depicting IgA, IgM and dual IgA and IgM targets for IgA^+^IgM^-^, IgA^-^IgM^+^ and IgA^+^IgM^+^ populations. **E.** Scatterplot of the IgM and IgA coating probability for each amplicon sequence variant (ASV) (n=107). Symbols with horizontal or vertical error bars are enriched in IgM or IgA binding (FDR<0.05). Error bars depict standard error of the mean. **F.** Percentage of fecal microbes coated by serum IgG and IgM from healthy pediatric subjects. **G.** Binding probabilities of representative IgG target microbes. **H.** Scatterplot of the IgG and IgM coating probability for each ASV (n=107).

To explore the relationship between microbes bound by sIgM and sIgA, we applied microbial flow cytometry and 16s rRNA gene sequencing (mFLOW-Seq)(Fig 1A) and sorted four populations of microbes based on the presence or absence of coating with sIgA and/or sIgM [IgM only (IgM^+^IgA^-^), IgA only (IgM^-^IgA^+^), dually coated with IgA and IgM (IgM^+^IgA^+^) or coated by neither IgM nor IgA (IgM^-^IgA^-^)] by fluorescence-activated flow sorting. (**Fig S1B**). To enhance purity, we performed a secondary sort on each population (Fig 1C). DNA was then extracted from each of these purified populations, and microbiomes were assessed by sequencing amplicons from the V4 region of the 16s rRNA gene. In parallel, we sequenced the same region of the 16s rRNA gene from the unsorted fecal microbiome for each subject. 5 million amplicons were analyzed from 199 sorted populations (median 20,000 reads per sample) and 0.5 million amplicons from the 32 unsorted fecal microbiomes (median 18,000 reads per sample). To obtain the most granular picture of the sorted and unsorted fecal microbiomes from the 16s rRNA gene sequences, we focused on the microbes defined as amplicon sequence variants (ASVs) present in at least a third of the fecal samples (N=107). The unsorted fecal microbiomes were dominated by microbes from five phyla: Firmicutes (N=81 ASVs, 56% relative abundance), Bacteroidetes (N=19, 33%), Actinobacteria (N=3, 3%), Proteobacteria (N=3, 4%), and Verrucomicrobia (N=1, 4%). The fecal microbiomes of healthy and SIgAD subjects were indistinguishable by either alpha or beta diversities or by comparison of the relative abundances of each microbial taxon (**Fig S1C-E).**

Weighted and unweighted UniFrac distances(Lozupone et al., 2011) revealed distinct community structure of the four populations of antibody-coated microbes (IgM^+^IgA^-^, IgM^+^IgA^+^, IgM^-^IgA^+^ and IgM^-^IgA^-^) from IgA sufficient siblings (**Table S2**). Alpha diversity, as measured by Shannon diversity, was similar for the IgM^-^IgA^-^, IgM^+^IgA^-^ (“IgM positive”) and IgA^+^IgM^+^ (“double positive”) population, while the IgM^-^IgA^+^ (“IgA positive”) bound microbial community had lower alpha diversity (**Fig S1F**), indicating that fewer microbial taxa were present in the IgA sufficient siblings.

To explore the different models (“discrete” vs. “continuous”) of antibody binding to commensals in the gut, we employed Bayesian analysis to define the absolute probability on a scale of 0-100% that a microbe from a given ASV is bound by sIgA, sIgM or both(Jackson et al., 2021). The Bayesian approach provides several important advantages compared to other methods including avoiding artifactual associations based on different relative abundances of ASVs, increased power to detect differences in antibody binding to specific ASVs between experimental groups and lower coefficients of variation(Jackson et al., 2021). Linear mixed effect (LME) modeling was then applied to binding probabilities to define which ASVs are preferentially coated by sIgM or sIgA. We identified 8 ASVs highly coated by sIgA and 15 ASVs that were preferentially coated by sIgM with FDR<0.05 (Fig 1D and **S2A-B**). 8 ASVs were enriched in the IgA^+^IgM^-^ single positive fraction from IgA sufficient siblings, which included 6 Firmicutes with 3 ASVs from the genus *Ruminococcus*, 1 from the genus *Clostridium*, 2 from the family *Ruminococaceae*, and 2 from the phylum Bacteroidetes with 1 ASV from the genus *Bacteroides* and 1 from the genus *Alistipes* (Fig 1D and **S2A-B**).

The 15 ASVs enriched in the IgM^+^IgA^-^ single positive fraction from IgA sufficient siblings included 14 taxa from the phylum Firmicutes and 1 Proteobacteria from the genus *Sutterella*. 8 of the 14 of the Firmicutes bound by IgM^+^IgA^-^ were from the family *Lachnospiraceae* with 4 ASVs from the genus *Coprococcus* and 2 ASVs from the genus *Blautia*. The other taxa bound by sIgM included 2 ASVs from the family Clostridiaceae, 2 from *Ruminococaceae* and from 1 *Eubacteriaceae*. In total, we identified 23 ASVs mainly from the phylum Firmicutes that were preferentially bound by either sIgA (N=8) or sIgM (N=15) (Fig 1D and **S2A-B)**.

Since 10.7% of fecal microbes were coated by both IgA and IgM in healthy controls (**Fig S1A**), we applied LME analyses to define the taxa that are preferentially bound by IgA^+^IgM^+^ (“double positive”) compared to either IgM^+^IgA^-^ or IgM^-^IgA^+^. A total of 33 unique ASVs were enriched in the dually-coated IgA^+^IgM^+^ population (Fig 1D and **S2C**). This dually coated group of microbes was predominantly composed of Firmicutes but also encompassed a more phylogenetically diverse group of microbes than the IgA or IgM single positive microbiota. The dually coated IgA^+^IgM^+^ fraction found in the fecal microbiomes of healthy children included microbes from all 5 phyla. Of these 33 dually-coated microbes, 26 were Firmicutes, 3 Bacteroidetes, 2 Proteobacteria, 1 Verrucomicrobia, and 1 Actinobacteria. Notably, dually-coated microbes also include *Enterobacteriaceae*, *Bifidobacteria* and *Akkermansia* (Fig 1D and **S2C-D)**.

To integrate and visualize the binding probabilities of these sorted populations, we plotted the probability of sIgM or sIgA coating for each ASV, calculated by summing the probability for the single positive fractions plus the probability of the sIgA and sIgM double positive population (Fig 1E). Microbes to the left of the diagonal were preferentially bound by sIgM while those to the right of the diagonal are more likely to be bound by sIgA. ASVs found along the diagonal are evenly coated with sIgA and sIgM. Few microbes had a coating probability of less than 25% or greater than 75% for either IgA or IgM. Dually-coated microbes were present at higher than expected frequencies based on the single IgA and IgM binding probabilities (Fig 1D and **S2C-D**). This high frequency of dual-coating supports a model in which IgA and IgM bind cooperatively to a diverse and mostly shared set of microbes.

In summary, within the most abundant phyla, microbes from Firmicutes taxa were more likely to be coated by sIgA and/or sIgM than Bacteroidetes. Further, ASVs from the less abundant phyla Proteobacteria (*Enterobacteriaceae*, *Sutterella*), Actinobacteria (*Bifidobacteria*) and Verrucomicrobia (*Akkermansia*) were highly coated by both sIgM and sIgA. Finally, the distribution of binding probabilities supports a model in which the probability of sIgA and sIgM coating is continuous for most microbes while a few taxa such as *Coprococcus* and *Ruminococcus* are discretely targeted by IgA or IgM, respectively (Fig 1D-E).

### Systemic IgM and IgG bind an overlapping subset of commensal microbes enriched in Firmicutes, Actinobacteria and Proteobacteria

Recently, systemic IgGs capable of binding to commensal enteric microbes were identified in healthy people(Fadlallah et al., 2019) and mice(Koch et al., 2016; Zeng et al., 2016). Yet, the degree to which anti-commensal antibodies are present in children, the specific commensal microbes targeted by systemic IgGs, the relationship between the mucosal and systemic humoral anti-commensal networks and the implications of these IgG for systemic immune reactivity remain poorly understood. To determine the degree of binding of serum IgGs to commensal fecal microbes in children we incubated fecal microbes with autologous serum (which contains IgA, IgM and IgG) to allow binding of systemic antibodies to fecal microbes each IgA sufficient subject’s serum with their own fecal microbiota as part of the mFLOW-SEQ workflow (Fig 1A), which revealed that systemic IgG antibodies bind to 7.6% of fecal microbes, most of which were dually coated with IgG and IgM (Fig 1F). Further, the addition of serum IgM led to an increased proportion of microbes bound by IgM from 25.8% to 47.6% (comparing Fig 1B and 1F). Thus, a substantial proportion of fecal microbiota are capable of being bound by circulating IgG and IgM antibodies in healthy children.

To define the commensal microbial targets of systemic IgG and IgM, we analyzed the microbiomes of 4 populations of antibody-bound microbes [IgM only (IgM^+^IgG^-^), IgG only (IgM^-^IgG^+^), dually coated with IgM and IgG (IgM^+^IgG^+^) and coated by neither IgM nor IgG (IgM^-^IgG^-^)] (Fig 1A and **S1B**). Alpha and beta diversities differed between the 4 IgG/IgM populations indicating that these sorted fractions represent distinct components of the microbiome (**Table S2** and **Fig S1G**). 5 ASVs from the fecal microbiome were highly targeted by systemic IgG. These 5 IgG targeted microbes belong to the phylum Firmicutes and were distributed across 1 order Clostridiales and 3 families (*Peptostreptococcaceae*, *Lachnospiraceae* and *Ruminococcaceae)* and 1 genus (*Ruminococcus*) (Fig 1G and **S3A**). IgM targeted 39 ASVs preferentially, including 35 Firmicutes, 2 Proteobacteria, 1 Actinobacteria and 1 Verrucomicrobia, while none of the Bacteroidetes ASVs were preferentially bound by serum IgM (**Fig S3B**). 29 ASVs were dually targeted by IgG and IgM (**Fig S3C-D**).

To integrate and visualize the binding probabilities across the different sorted populations, we plotted the probability of IgM and IgG coating for each ASV, calculated by summing the probability for the single positive fractions plus probability of the double positive (e.g., IgM^+^ IgG^+^) fraction (Fig 1H). ASVs to the left of the diagonal were preferentially bound by IgM, and ASVs found along the diagonal were evenly coated with IgM and IgG. No sequenced microbe had a coating probability of less than 25% for IgM, and 41 of 107 ASVs had a greater than 75% coating probability for IgM. All microbes exhibited a higher probability of IgM than IgG binding.

We next asked how systemic IgG and mucosal IgA and IgM anti-commensal antibody networks relate to each other by comparing the binding probabilities of each antibody isotype to each ASV. First, IgM and IgA binding probabilities were correlated across the 107 ASVs (Spearman r=0.49, p=7.3 x 10^-8^; Fig 2A) indicating that these two main secretory isotypes bind to common microbial targets. Second, the probability of systemic IgG coating was highly correlated with the likelihood of fecal sIgA coating the same ASV (Spearman r=0.66, p=6.3 x 10^-15^) (Fig 2B). Similarly, fecal sIgM binding also correlated with the probability of serum IgG coating (Spearman r=0.62, p=1.2 x 10^-12^) (Fig 2C). Further, the 5 most IgG-targeted microbes were also highly coated by sIgM and sIgA (binding probability range, 55-74% for IgM and 57-85% for IgA) (**Table S3**). All 5 IgG-targeted microbes were within the top 20 most IgA and IgM coated microbes and included 4 of the 8 most highly IgA coated taxa. In addition, there were almost complete overlap of microbes with greater than 50% binding probability for either sIgM, sIgA or IgG (Fig 2D-E). 28 of the 32 ASVs targeted by IgG were also bound by IgA and/or IgM (Fig 2D-E). These strong correlations between the main mucosal antibody isotypes sIgA and sIgM, and systemic IgG targets, demonstrated that the mucosal and systemic antibody networks target a broad range of microbes to a similar degree and suggested connections between these antibody isotypes.

**Figure 2.**
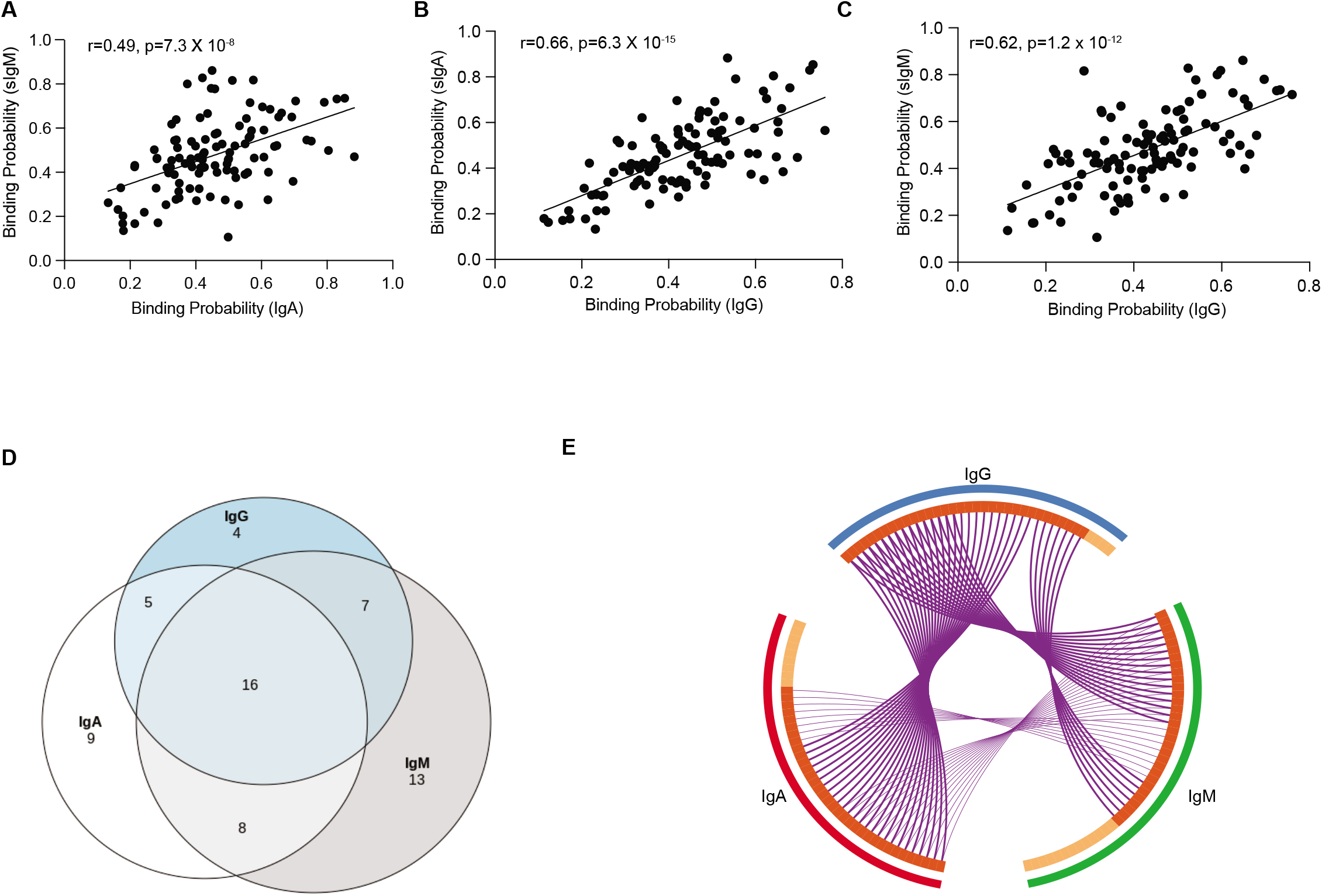
Mucosal and systemic antibodies target a common set of fecal microbes in healthy children. **A-C.** Correlation of the binding probability to each ASV by secretory IgM, IgA and serum IgG. Spearman correlation, line represents simple linear regression. **D.** Venn diagram and **E.** Circos plot depicting the relationship of microbial ASVs with a binding probability >50% in either IgA, IgM or IgG. Purple lines connect ASVs bound >50% for both of the indicated antibody isotypes. Overlapping bacterial ASVs are shown by the red part of inner ring (IgG n=28, IgA n=29 and IgM n=31). Bacterial ASVs bound by only one isotype are represented by the orange part of inner ring (IgG n=4, IgA n=9 and IgM n=13).

### Fecal and serum IgA discordance

Our recruitment strategy identified SIgAD patients and unaffected siblings. Many of the healthy siblings had not previously had IgA levels checked in our system and unexpectedly 4 siblings enrolled as healthy controls had undetectable levels of serum IgA (<7mg/dL; **Fig S4A**). These 4 siblings met the clinical definition for SIgAD, though they were not previously diagnosed (**Table S4**). In addition, we identified discordance between serum and fecal IgA in a subset of healthy siblings and SIgAD patients. Specifically, free and bacteria-bound fecal IgA was detected in 5 of the 19 SIgAD subjects (free fecal IgA range, 26.4-123.2 µg/g, microbe-bound fecal IgA range 0.9-16.7 µg/g) and confirmed by microbial flow cytometry in these subjects (**Fig S4B-D** and **Table S4**). Similarly, a subset of 6 SIgAD subjects had detectable IgA^+^ memory B cells in circulation (**Fig S4E**). Conversely, one control subject had normal serum IgA concentration (101 mg/dL) but lacked fecal IgA. Since serum IgA concentration and fecal IgA levels were discordant in 6 of 32 study subjects, we defined each subject’s IgA status, based on the presence and binding of IgA to commensal microbes, as either “fecal IgA positive” or “fecal IgA negative”. We used stringent criteria for fecal IgA negative which required less than 10 ug/g free fecal IgA, less than 1 ug/g bacteria bound sIgA and less than 5% IgA bound fecal microbes by microbial flow cytometry (**Table S4**). Applying these criteria to this cohort yielded 15 fecal IgA negative and 17 fecal IgA positive pediatric subjects (**Fig S4F)**. Using a fecal IgA definition, rather than a serum definition, allows a more rigorous investigation of the impact of intestinal IgA on the fecal microbiome and systemic immune system. In this cohort, serum IgA concentration did not correlate with free fecal IgA concentration, quantity of IgA bound to fecal microbes, or percentage of fecal microbes coated by IgA with serum IgA levels (**Fig S4G**), which supports using fecal IgA parameters rather than serum IgA concentration to define the mucosal IgA status of subjects for our study.

### sIgM only minimally compensates for the absence of intestinal sIgA

Since sIgM is often invoked as a compensatory molecule for the functions of IgA in subjects lacking IgA(Catanzaro et al., 2019; Fadlallah et al., 2018; Macpherson et al., 2018), we investigated several potential mechanisms by which sIgM may respond to IgA deficiency. We observed similar fecal concentrations of IgM and proportions of fecal microbes coated with sIgM in fecal IgA deficient and sufficient subjects (Fig 3A-B). Based on these findings, we conclude that fecal IgA deficient subjects in this cohort did not compensate for the absence of fecal IgA by either increasing total fecal IgM concentration or by having IgM capable of binding to a higher proportion of fecal microbes.

**Figure 3.**
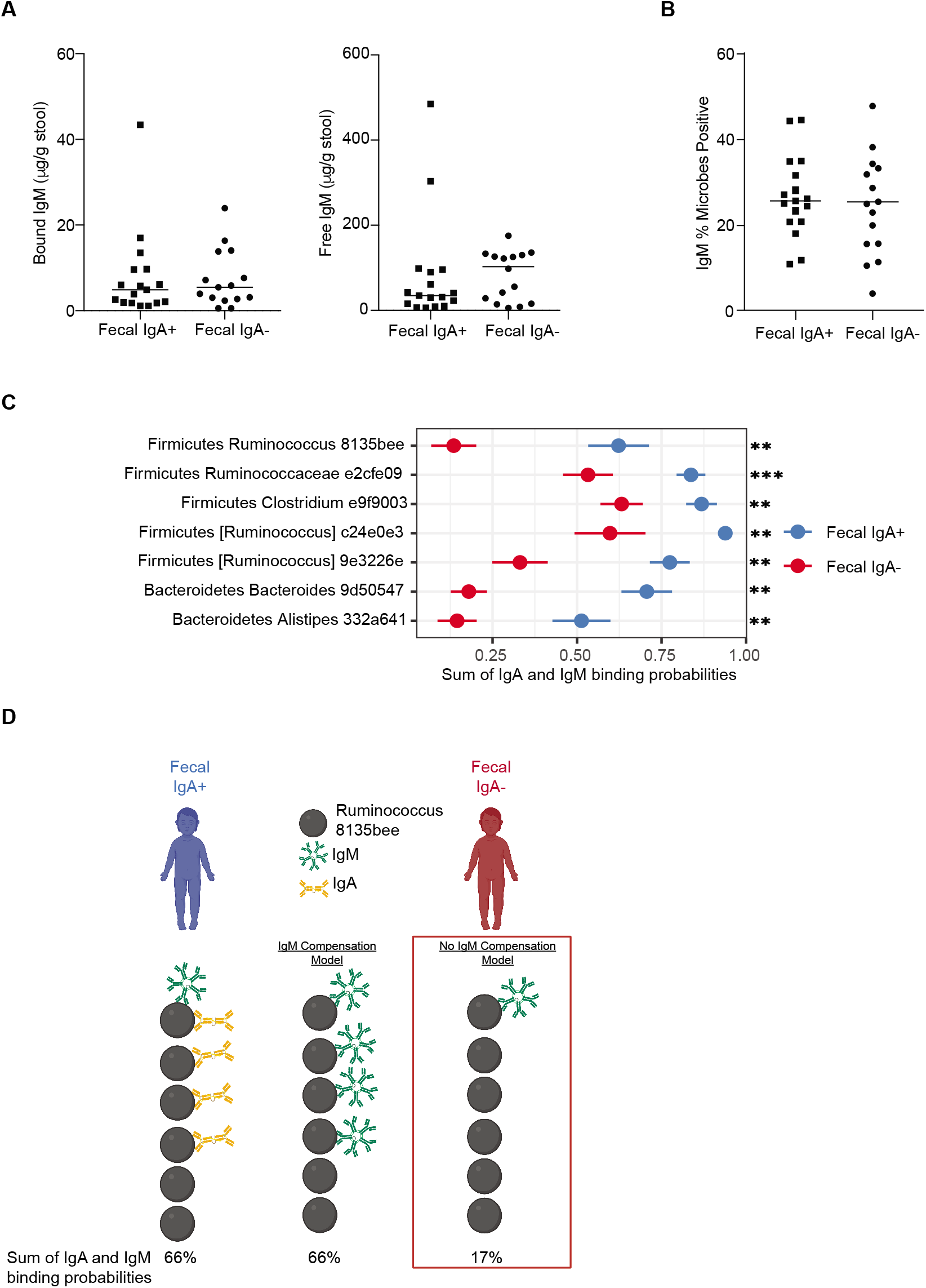
Limited compensation by secretory IgM in subjects with fecal IgA deficiency. **A.** Bacteria-bound and free IgM concentration from stool samples of fecal IgA+ (n=17) and fecal IgA-subjects (n=15) measured by ELISA. **B.** Percentage of fecal microbes bound by IgM in subjects with fecal IgA deficiency and IgA+ control siblings determined by microbial flow cytometry. **C.** Comparison of the total binding probabilities of fecal microbes coated by secretory immunoglobulins (IgA plus IgM) in subjects lacking fecal IgA (red) and control siblings (blue). Bars represent SEM. ** p<0.01 and ***P<0.001. **D.** Schematic of secretory IgM and IgA binding probabilities in controls and fecal IgA-subjects.

To address whether sIgM compensates for the lack of fecal IgA by coating those microbes which are normally coated with IgA, we double-sorted IgM-uncoated and IgM-coated microbial populations from fecal IgA deficient and sufficient siblings. 16s rRNA gene-based microbiome analysis of these sorted population revealed that IgM-coated microbes from IgA deficient subjects had similar Shannon diversity as the IgA^+^IgM^+^ coated microbial population and slightly higher diversity than the IgA^-^IgM^+^ population from sibling controls. This modest increase in alpha diversity suggests that IgM antibodies in IgA deficient subjects coat additional taxa compared to IgM from healthy siblings. Weighted and unweighted UniFrac distances were similar between IgM-coated and IgM-uncoated microbial communities from healthy and IgA deficient subjects. Overall, this analysis provides evidence that microbes bound by IgM are quite similar between IgA deficient and IgA sufficient siblings. In addition, if IgM binding to commensal microbes did compensate for IgA deficiency, we anticipated that IgM binding would increase for those commensals coated by IgA in healthy subjects. The probability of IgM binding to the top 7 IgA targeted microbes remained significantly lower in IgA deficient subjects relative to the total probability of binding to IgA or IgM for the same ASVs in healthy sibling (Fig 3C-D). This lack of compensatory IgM binding to microbes normally highly coated by IgA argues that sIgM does not compensate for IgA deficiency by enhanced binding to IgA targeted microbes. In summary, pediatric subjects with fecal IgA deficiency in our cohort did not increase fecal IgM (Fig 3A-B), nor did their sIgM repertoire quantitatively adjust to target those microbes typically targeted by sIgA (Fig 3C-D). Taken together, we find limited evidence for IgM compensating quantitatively or qualitatively in pediatric patients lacking sIgA.

### Serum IgGs bind to a higher proportion of fecal microbes in subjects lacking fecal sIgA

Since sIgM did not effectively compensate for sIgA in pediatric subjects lacking fecal IgA, we next investigated the impact of IgA deficiency on the predominant circulating antibody isotypes, IgM (present in both compartments) and IgG (present in periphery alone). We hypothesized that sIgA restrains systemic exposure to commensal microbes, and that subjects lacking sIgA would therefore have increased systemic exposure to commensal microbes and a compensatory increase in systemic antibody responses to commensal microbiota. Quantitatively, serum IgM concentrations and the proportion of circulating IgM^+^ memory (CD27+ CD19+) B cells were similar between subjects lacking fecal IgA and their control siblings (**Fig S5A-B**). In contrast, subjects lacking fecal IgA had 41% higher serum IgG concentration (median 1,280 mg/dL vs 907 mg/dL, p=0.02) and increased proportion of circulating IgG^+^ IgA^-^ memory (CD27+ CD19+) B cells (Fig 4A). These findings demonstrate that fecal IgA deficiency impacts the systemic humoral compartment.

**Figure 4.**
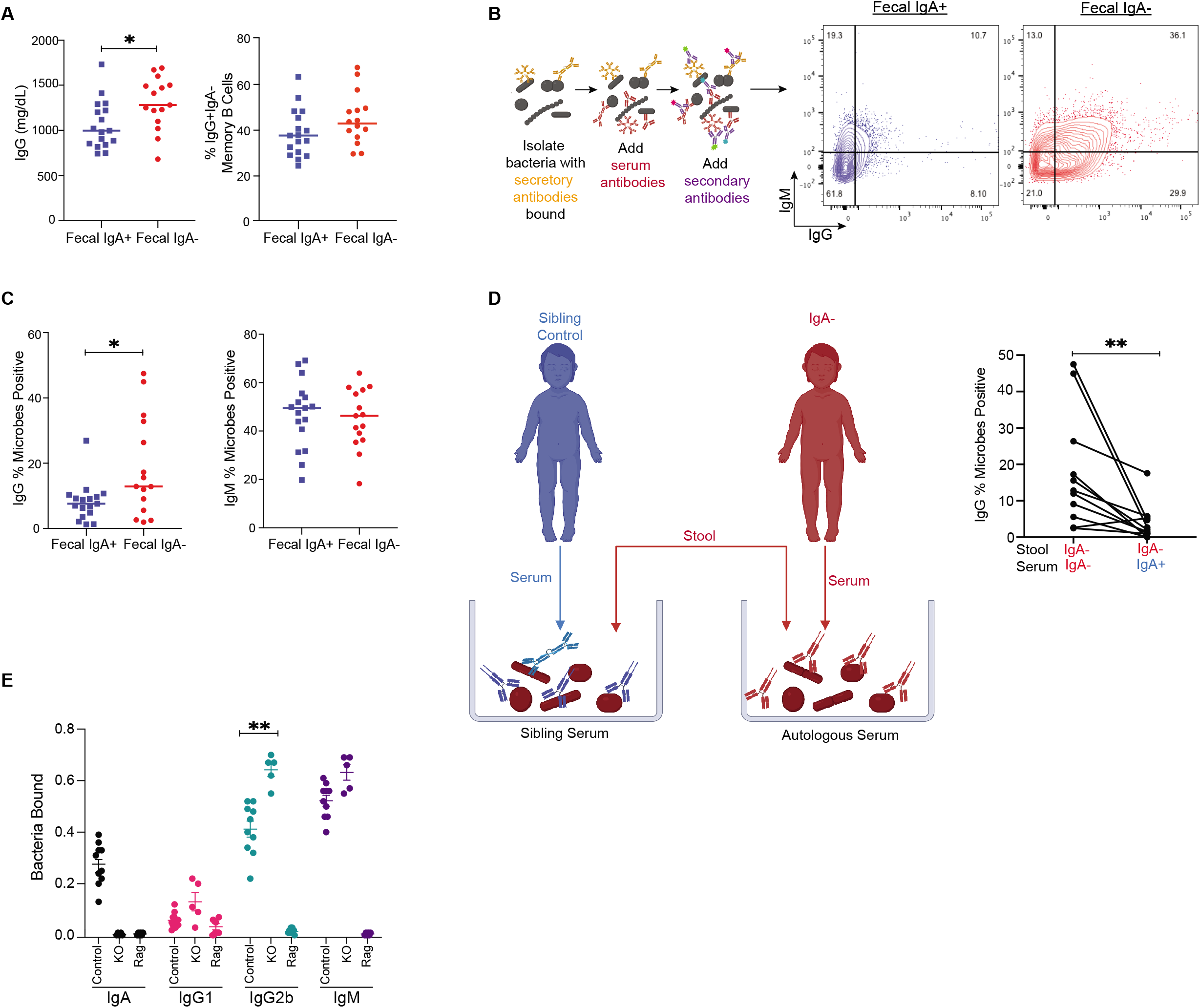
IgA restrains systemic targeting of commensal microbes. **A.** Serum IgG concentration and percentage of circulating IgG+ memory B cells. **B.** Microbial flow cytometry (mFLOW) schematic and gating scheme. Fecal microbes were incubated with autologous serum (normalized to 10 ug/mL of IgG) stained with the nucleic acid dye Syto BC and either fluorophore conjugated anti-IgM or anti-IgG antibodies or isotype controls and analyzed by flow cytometry. mFLOW plots from fecal IgA+ subject (left) and fecal IgA-subject (right). **C.** Percentage of fecal microbes coated by IgM or serum IgG**. D. Left:** Schematic of serum swap experiment between fecal IgA+ (blue) and fecal IgA-(red) siblings. Fecal microbes from fecal IgA-subjects were incubated either with autologous (fecal IgA-) or sibling (fecal IgA+) serum and analyzed by flow cytometry. **Right:** Percent of fecal microbes from fecal IgA-subjects bound by IgG in autologous or sibling serum. **E.** Microbial flow cytometry of the proportion of fecal microbes from IgA deficiency mice (KO) or IgA heterozygous (Control) littermates coated with IgA, IgM, IgG1 or IgG2b. Serum and stool from RAG2 deficient mice which lack antibodies were used as negative controls. Data shown are representative of results from 4 independent experiments. *P<0.05, **P<0.01.

We next considered whether an increased systemic IgG response against commensal microbes accounted for the increase in serum IgG concentration. To test this hypothesis, we incubated fecal microbes from each subject with autologous serum (normalized to a concentration of 10 ug/mL of total IgG) to determine the proportion of fecal microbes targeted by systemic IgG and IgM antibodies. Circulating IgG antibodies from subjects lacking fecal IgA coated a significantly higher proportion of their own intestinal commensal microbes than sibling controls (median 12.9% vs 7.6%, p<0.05) (Fig 4B-C and **S5C-D**). This enhanced binding was found across a range of IgG concentrations (**Fig S5E**). In contrast, serum IgM binding to commensal microbes was indistinguishable between healthy and IgA deficiency siblings (median 49.5% vs 46.3% p=0.68) (Fig 4B-C).

Since some commensal microbes bind to host antibodies, through interactions with the crystallizable fragment (Fc) portion of IgG (e.g., protein A and G or B cell superantigens)(Bunker et al., 2019), we compared binding of serum antibodies from each fecal IgA deficient and sufficient sibling pair to the same fecal microbiota. Serum IgG from IgA deficient sibling coated a higher proportion of autologous fecal microbes than serum IgG from IgA sufficient sibling (Fig 4D). Therefore, the serum IgG antibodies of IgA deficient subjects have increased binding capacity to autologous commensals compared to IgA sufficient siblings.

Similarly, in a murine model of IgA deficiency(Harriman et al., 1999), serum IgG2b from IgA deficient mice coated a significantly higher proportion of fecal microbes compared to littermate control mice (Fig 4E). The enhanced systemic IgG response was seen shortly after weaning in mice 5-6 weeks of age which corresponds to early induction of anti-commensal IgG2b in mice(Koch et al., 2016), indicating a non-redundant role for IgA in restraining systemic responses to commensal microbes early in immune system development. We observed increased anti-commensal IgG binding from IgA deficient mice versus littermate controls when comparing binding to fecal microbes from either IgA deficient mice or littermate controls (**Fig S5F**). Taken together, these findings in humans and mice demonstrated that sIgA restrains the total quantity and microbial specificities of circulating IgG antibodies.

Since serum IgG bound to a higher proportion of commensal microbes in IgA deficient siblings, we explored whether specific microbes were targeted by systemic IgGs across subjects or whether IgG targeting was private for each individual as has been suggested (Fadlallah et al., 2019). To define the commensal microbial targets of systemic IgG and IgM in subjects with IgA deficiency and their healthy siblings, we double-sorted and sequenced the microbiomes of 4 populations of antibody-bound microbes [IgM only (IgM^+^IgG^-^), IgG only (IgM^-^IgG^+^), dually coated with IgM and IgG (IgM^+^IgG^+^) and coated by neither IgM nor IgG (IgM^-^IgG^-^)]. Alpha and beta diversities differed between sorted populations of IgM and IgG coated microbes (Fig 5A and **Table S2**). The IgM^-^IgG^+^ microbiomes had the lowest diversities in fecal IgA+ and fecal IgA-subjects, which suggests that a more limited set of microbes are bound by the IgG than IgM. Alpha and beta diversities were similar between fecal IgA+ and fecal IgA-subjects. The probabilities of IgM and IgG binding were broadly similar between fecal IgA+ and IgA-siblings with a modest non-significant shift of some taxa towards higher IgG probability in the IgA deficient group (Fig 5B-C). Yet, in a subset of fecal IgA deficient subjects who have highly elevated IgG binding to commensals (range 17-47%), we found a high binding probability for their IgG antibodies to the two most highly abundant Bacteroidetes ASVs and to *Enterobacteriaceae* which notably included microbes with pathogenic potential (e.g., *E. coli*, *Klebsiella,* and *Enterobacter)* (Fig 5C). In subjects with fecal IgA deficiency, total fecal IgG binding was correlated with the probability of IgG binding to *Enterobacteriaceae* which suggests that increased IgG binding is driven in part by microbes with the potential for translocation and systemic infection (Fig 5D). These findings raise the possibility that in the absence of secretory IgA, a compensatory increase of circulating IgG antibodies specific for commensal microbes helps maintain homeostatic immunity. However, given that IgA deficiency is a disease of affected outliers, in that only a subset of patients who lack serum IgA are symptomatic, it was necessary to delve deeper into the baseline immune state of SIgAD.

**Figure 5.**
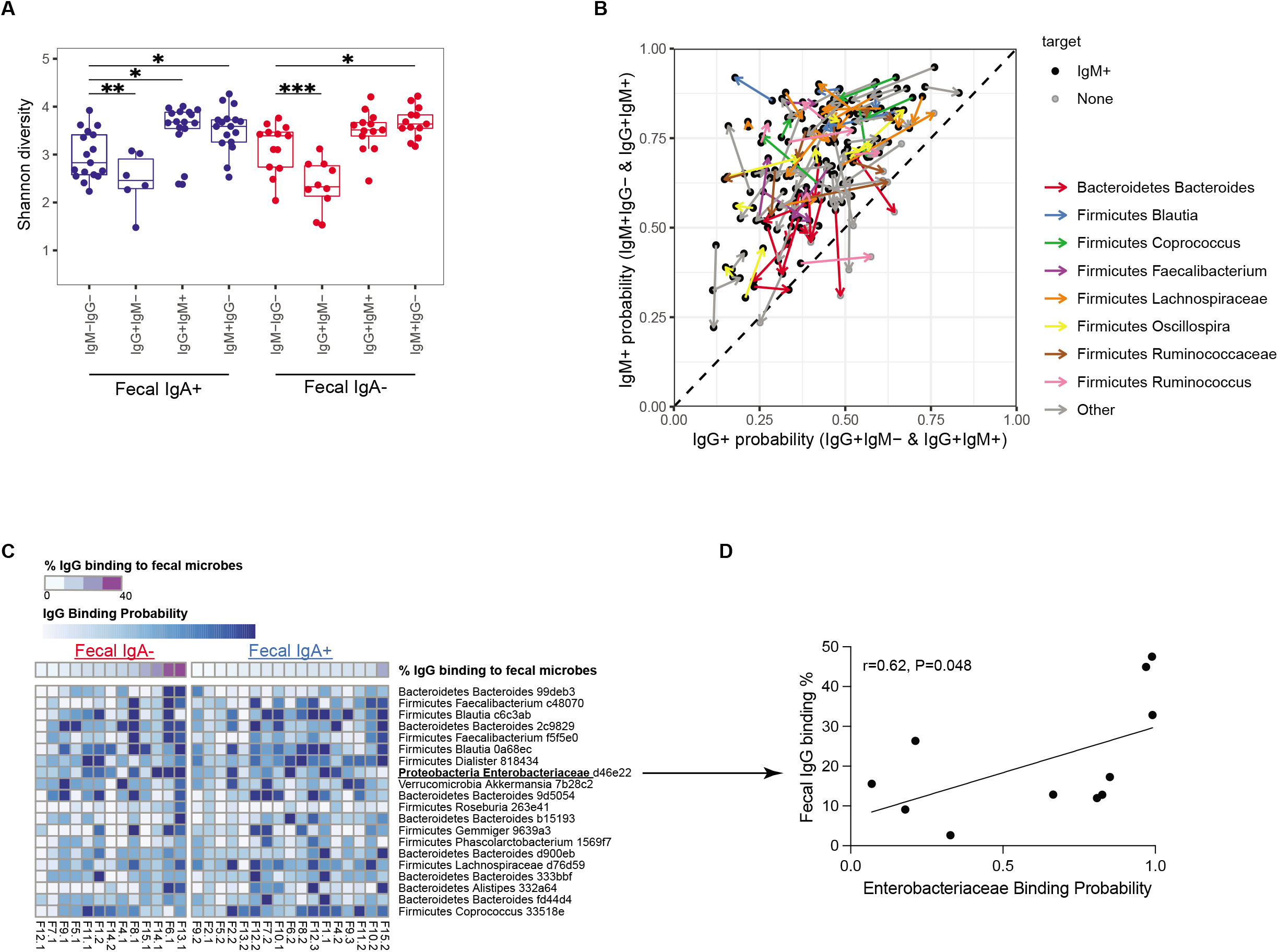
IgG targeting of commensal microbiota in subjects lacking fecal IgA. **A.** Alpha diversities of the sorted fecal microbiomes. **B.** Scatterplot of the IgM and IgG coating probability for each ASV (n=107). Each dot represents a single ASV, and arrows connect average IgM and IgG binding probabilities starting from control and ending with IgA deficient subjects. **C.** IgG binding heatmap with each individual on the x axis and the top 20 most abundant fecal microbes on the x axis. D. Fecal IgG binding percentage with probability of IgG coating Enterobacteriaceae in subjects with fecal IgA deficiency (n=11). Spearman correlation, line represents simple linear regression. *P<0.05, **P<0.01, ***P<0.001.

### Systemic immune dysregulation in pediatric subjects lacking intestinal IgA

To more deeply interrogate the immunophenotype in fecal IgA deficiency, we used Cytometry by Time Of Flight (CyTOF)(Bendall et al., 2012) of PBMCs from the cohort to define the high dimensional immune profile in subjects with fecal IgA deficiency versus that of IgA sufficient subjects, paired with flow cytometric analysis of peripheral (splenic) and local secondary lymphoid tissue (MLN) immune cell populations from IgA deficient and littermate control mice.

First, peripheral blood CD4 T helper subsets, as defined by surface markers and canonical transcription factors, were found to be unaltered in frequency in IgA deficient humans or mice (**Fig S6A** and data not shown). We next defined T cell activation states in human CD4 and CD8 T cells using CyTOF of PBMCs with manual gating and a dimensionality reduction algorithm, Phenograph, to cluster live cells (Fig 6A)(Chen et al., 2016; Levine et al., 2015). Therein we identified one cluster of effector memory CD8 T cells (i.e., cluster 15, with elevated levels of: Helios, TIGIT, Eomes, CD39) that was significantly increased in fecal IgA deficient subjects (Fig 6A). Since cells from this cluster expressed multiple inhibitory receptors, we investigated the presence of cellular immune dysregulation by investigating all clusters with altered levels of multiple T cell activation and inhibitory markers. We focused on a subset of T cell clusters that, like cluster 15, had overexpression the expression of several inhibitory receptors and/or cell surface markers that can be seen in T cell exhaustion, or systemic immune dysregulation more broadly (**Fig S6B**). Using an outlier analysis with a permutation p-value threshold of 0.05 and a Fisher’s exact test, we found that there was a non-significant enrichment of overexpression a subset of T cell exhausted-like clusters in fecal IgA deficient subjects (data not shown).

**Figure 6.**
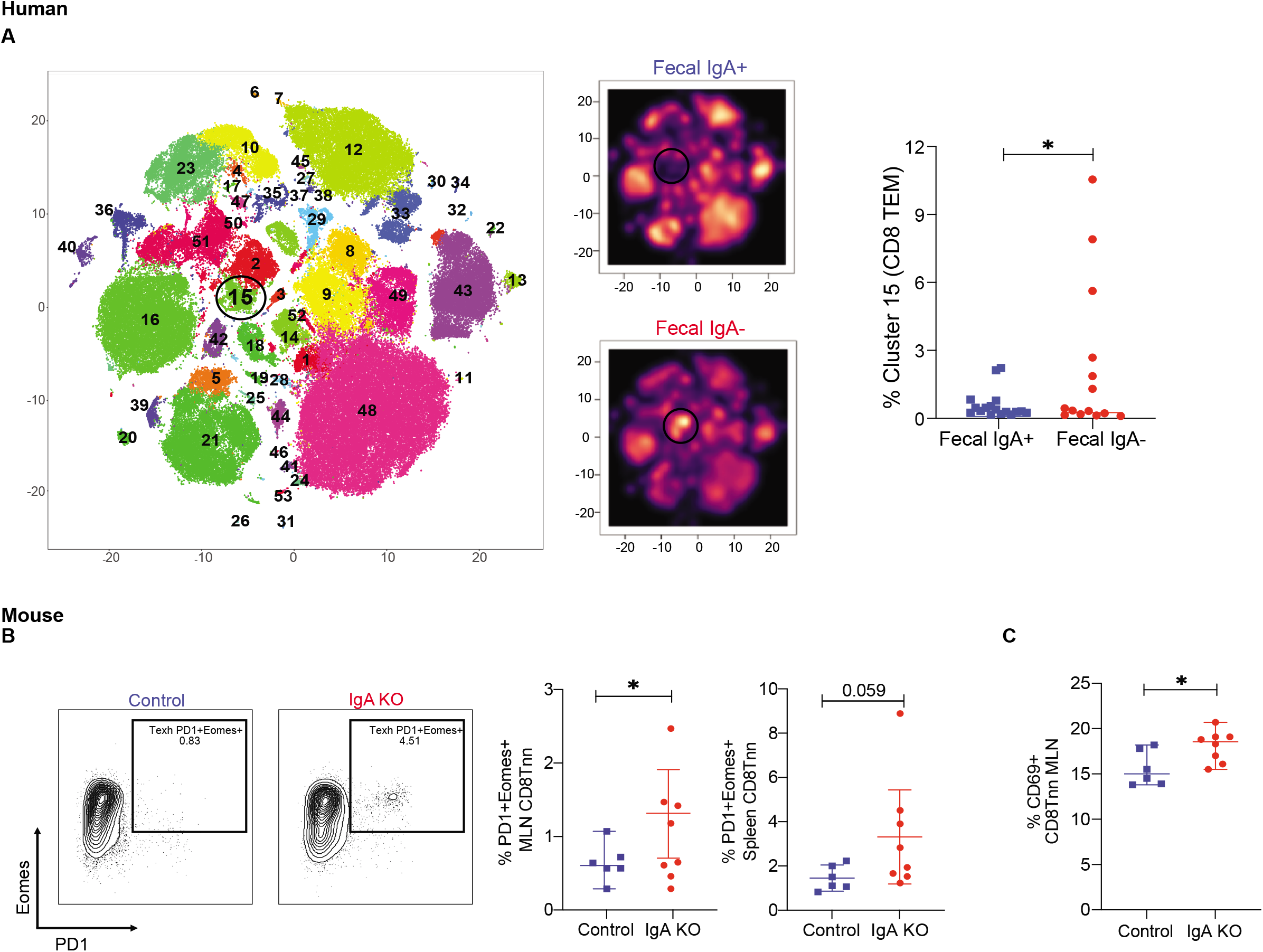
Cellular immune dysregulation in subjects lacking fecal IgA. **A.** Immunoprofiling. PBMCs were labeled with metal-conjugated antibodies and detected by mass cytometry (CyTOF). Dimensionality reduction was performed using Rphenograph to cluster 5,000 live cells per subject, with cluster diagram as shown on the left and galaxy plots shown to the right. Black circle highlights cluster 15 that is enriched in the fecal IgA+ group. Scale: black or purple indicates fewer cells, yellow indicates more cells in each cluster. Scatterplot showing cluster participation frequency between fecal IgA+ and fecal IgA-subjects in the CD8 T exhausted cluster 15. **B.** PD-1 and Eomes expression in CD8 T cells from the spleens of IgA KO mice and littermate control IgA heterozygous mice with summary plots from the mesenteric lymph nodes (MLN) and spleen to the right. Data are representative of results from 2 independent experiments. *P<0.05. **C.** Percentage of CD8 T cells expressing CD69 in MLNs of IgA KO and littermate control IgA heterozygous mice.

Given our evidence of altered human T cell activation state, we further explored possible links to systemic immune dysregulation in a murine system by comparing T cell activation at baseline in IgA deficient and littermate control mice. Furthering the picture of altered CD8 T cell activation state in our human data, IgA deficient mice had an increased percentage of PD-1^+^ Eomes^+^ CD8 non-naïve T cells (Tnn) and CD69^+^ CD8 Tnn in the gut-draining mesenteric lymph nodes (MLN) compared to littermate control mice (Fig 6B-C). Given the altered immunophenotype, we examined the capacity of splenocytes to produce cytokines using a PMA/ionomycin stimulation to better assess T cell function and found no significant alterations in CD8 Tnn secretion of IFN-*γ,* though alterations in CD8 T cell cytokine secretion were identified (**Fig S6D)**. Thus, in both human and mouse IgA deficiency, we found evidence of altered CD8 T cell activation, consistent with systemic immune dysregulation, though testing the mechanistic link between IgA deficiency and immune dysregulation, and its potential implications for prognosis, will require further study and a larger cohort of subjects.

Since systemic immune responses, including vaccine responses, may be modulated by commensal microbes through a number of mechanisms including changes in gut permeability, metabolites and cytokine dysregulation(Lynn et al., 2021), we compared retained vaccine responses to *S. pneumoniae*, measles, mumps, diphtheria and tetanus in our pediatric cohort but found no significant differences (**Fig S6E**).

To more deeply investigate the immune systems of SIgAD fecal IgA deficient subjects, we measured key serum cytokines across our cohort. Out of 38 cytokines assayed, we identified 27, including type 1 (IFN-*γ*, IFN*α*2, IL-12), type 2 (IL-4 and IL-13) and type 17 (IL-17A) and inflammatory cytokines (e.g., IL-6 and TNF*α*) that were significantly increased in fecal IgA deficient subjects while only 1 (CXCL1) was significantly increased in the controls (Fig 7A and **S7A**), demonstrating dysregulation inflammation in our human fecal IgA subjects. In parallel, in murine IgA deficiency, we found a similar broad pattern of serum cytokine dysregulation with increased type 1, type 2 and pro-inflammatory cytokines (Fig 7B and **S7B)**.

**Figure 7.**
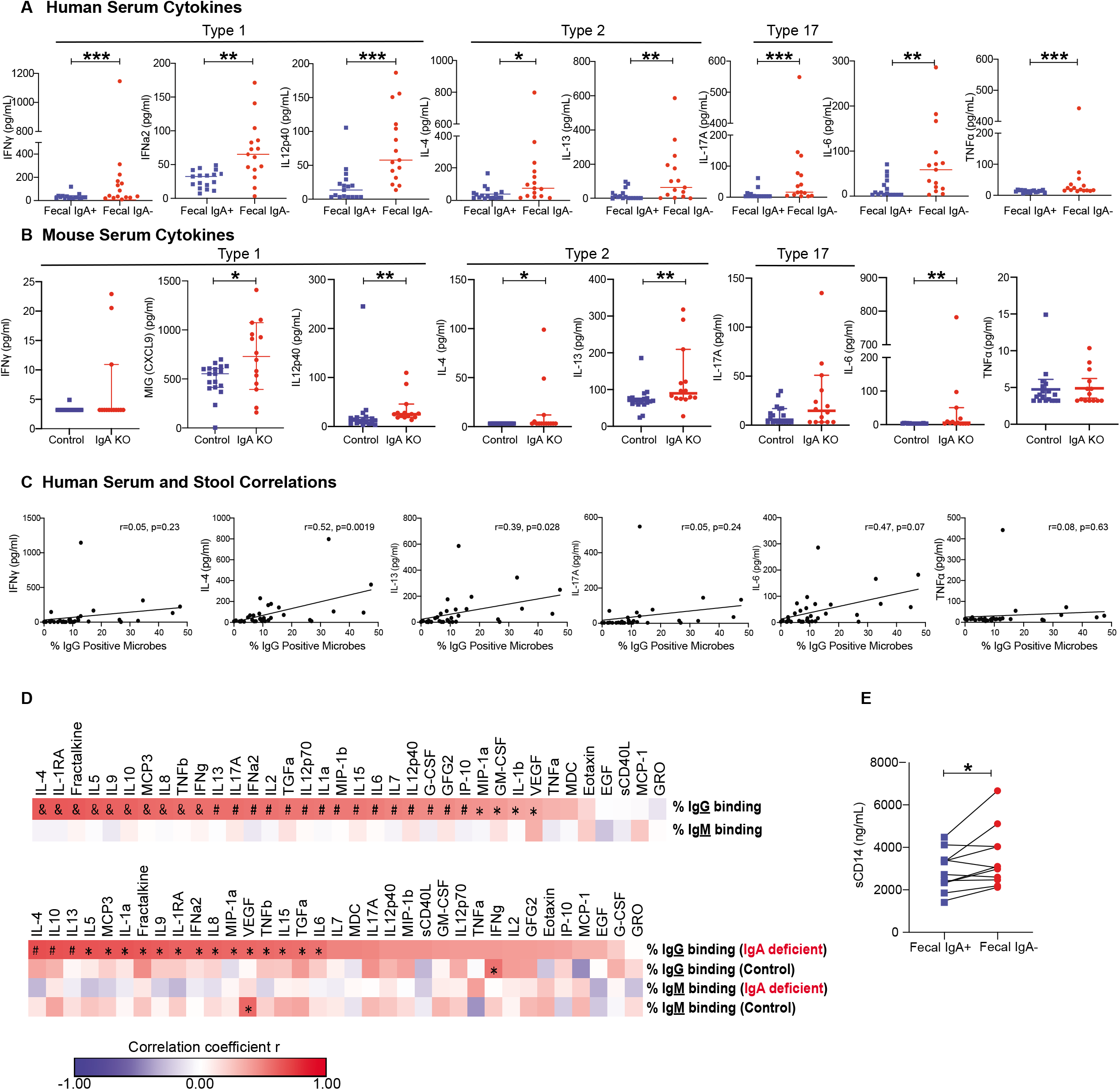
Humoral and cytokine dysregulation in subjects lacking fecal IgA. **A.** Serum cytokine concentrations in patients with fecal IgA deficiency (n=15) and control siblings (n=17). **B.** Serum cytokine concentrations (pg/ml) in IgA heterozygous control (n=18) and littermate IgA KO (n=14) mice. **C.** Spearman correlation of the percent of IgG binding to fecal commensal to serum cytokine concentration. Line is simple linear regression. **D.** (**Top)** Heatmap of the Spearman correlation coefficient of the percent of IgG and IgM binding. (**Bottom)** Heatmap of the correlation of the percent of IgG and IgM binding in IgA deficient subjects compared to sibling controls. * P<0.05, # P<0.01, & P<0.001. **E.** Soluble CD14 (ng/mL) in IgA deficient subjects compared to their healthy siblings. Paired t-test.

Having observed dysregulation of many cytokines in the fecal IgA deficient cohort, we next tested whether this was due to a few subjects with broadly dysregulated serum cytokines versus single or few cytokines dysregulated in each subject. Using an outlier analysis motivated by Fisher’s method of meta-analysis with a permutation p-value threshold of 0.05 (see Methods), we found a significant increase in pan-cytokine levels (broad elevation of serum cytokine expression) in a subset of fecal IgA negative subjects (**Fig S7C**). When assessed overall, there was a significant increase in pan-cytokine dysregulation (elevation of cytokine expression across all assessed cytokines) in fecal IgA negative subjects (p = 0.038; Fisher’s exact test) and a trend towards type 2 immunity-related (IL-4, IL-5 and IL-13) cytokine upregulation (p = 0.092; Fisher’s exact test, data not shown).

We next investigated whether there was a relationship between the level of systemic IgG binding to commensal microbes and systemic cytokine concentrations. IgG binding to commensals was correlated with type 1 (IFN-*γ*), type 2 (IL-4 and IL-13), type 17 (IL-17A) and other pro-inflammatory cytokines (IL-6) (Fig 7C). This correlation was driven by enhanced IgG binding in the subjects lacking fecal IgA (Fig 7D). In contrast, there were no statistically significant correlations between systemic cytokines and the proportion of IgM binding to commensals (Fig 7D). These findings demonstrate that systemic anti-commensal IgGs were specifically correlated with dysregulated cytokines in subjects lacking fecal IgA. Serum IL-4, IL-5, IL-10 and IL-13 were the most highly correlated cytokines with IgG binding in IgA deficient subjects (Fig 7D).

Given the presence of significant dysregulation of cytokine expression, we examined the potential link between IgA deficiency and intestinal barrier dysfunction. Intestinal barrier dysfunction leads to increased systemic inflammatory responses to commensal microbes in a number of diseases(Brenchley and Douek, 2012). We investigated the integrity of intestinal barrier function by measuring a widely used marker of intestinal barrier dysfunction and bacterial translocation, the serum concentration of soluble CD14 (sCD14)(Brenchley and Douek, 2012). We found that sCD14 concentration was elevated in subjects lacking fecal IgA compared to their household sibling control (Fig 7E), supporting the hypothesis that IgA restrains access of microbial products to systemic sites. Since bacterial products (e.g., lipopolysaccharide, LPS) activate circulating monocytes to release sCD14, we investigated levels of anti-LPS antibodies and found similar levels of systemic IgM and IgG against LPS in control and IgA deficiency siblings (**Fig S7D**). Further, we did not observe elevated levels of TLR4 agonists in the serum of IgA deficient subjects using an ultra-sensitive TLR4 bioassay (data not shown)(Thaiss et al., 2018).

## Discussion

A long-standing paradox in clinical immunology has been the ability of most patients lacking IgA to remain apparently healthy and our current inability as a field to predict which patients will be symptomatic with recurrent infections, allergy or autoimmunity. In this study we find that sIgA restrains the systemic IgG response to commensal microbes and blocks the development of systemic immune dysregulation of both circulating CD8 T cells and serum cytokines in both humans and mice. These effects are modest in some IgA deficient subjects and striking in others. The differences suggest that the variable phenotypes of IgA deficiency may reflect the degree to which mucosal barrier function is compromised as well as differences in systemic responses to aberrant exposure to commensal microbes. Thus, we propose that sIgA tunes the homeostatic dialogue between systemic immune compartment and commensal microbiota.

Here, we consider the clinical and immunologic phenotypes of IgA deficiency and the importance of sIgA through the lens of homeostatic immunity in which there is a constant active dialogue between commensal microbes and the mucosal immune system at barrier sites, and less intense and perhaps intermittent interactions between commensal microbes and the systemic immune compartments. First, our study and others demonstrate that IgA is a key regulator of both mucosal and systemic immunity(Fadlallah et al., 2018; Fadlallah et al., 2019; Wilmore et al., 2018). Intriguingly, co-housed littermate IgA deficient mice also display variable levels of cellular, humoral and systemic cytokine dysregulation which suggests that the functions of IgA are often, but not always, well-compensated by other aspects of the immune system. This framing may help understand, and perhaps predict in the future, the variable clinical phenotypes in patients with IgA deficiency. Second, IgA deficiency helps refine our understanding of the connections between the mucosal and systemic immune systems (i.e., circulating anti-commensal IgG antibodies) which are present during homeostasis and become more obvious in the setting of IgA deficiency.

A long-standing hypothesis in the field posits that secretory IgM (sIgM) compensates for the functions of IgA, thereby mitigating clinical disease in patients with selective IgA deficiency. Notably, patients with fecal IgA deficiency in our cohort do not possess increased secretory fecal or serum IgM concentrations, nor does IgM compensate for the lack of IgA by increased IgM binding to IgA targeted microbes. In addition, if IgM fully compensated for IgA’s functions in immune exclusion of microbes and microbial antigens from systemic sites, the systemic responses to commensal antigens should remain unchanged. Instead, subjects with fecal IgA deficiency in our study had an increased concentration of systemic IgG and increased concentrations of IgG antibodies that can directly bind to commensal microbes, demonstrating increased systemic humoral responses to gut commensal antigens. Thus, our data supports a model wherein IgM does not fully compensate for the functions of IgA in subjects with IgA deficiency and propose that systemic IgGs provide an important layer of compensation to help maintain homeostatic immunity in patients with IgA deficiency.

In both mice and humans with IgA deficiency, we see a small population of CD8 T cells with increased expression of activation and inhibitory receptors at baseline, raising the possibility of a role for T cell exhaustion in a subset of patients in this disease of chronic inflammation (with elevated systemic cytokines) and systemic recognition of commensal gut microbes, which could yield the signals needed for the development of T cell exhaustion. Classically, in the presence of chronic antigen signaling and inflammation (e.g. malignancy and chronic viral infection; McLane, Ann Rev Imm, 2019) T cells can become exhausted, with alterations in inhibitory and activation markers (e.g. PD-1), transcriptional networks and epigenetic state, as well as impaired effector function. This dysregulated T cell state is therapeutically relevant, as immune checkpoint blockade, wherein monoclonal antibodies target inhibitory receptor signaling and lead to improved anti-malignancy T cell function have been shown to improve outcomes in a number of malignancies. Here, in humans, we identified a significantly increased frequency of an exhausted-like CD8 T cell cluster in a subset of our fecal IgA deficient subjects. In mice, we see local (MLN) but not systemic (splenic) increases in PD-1^+^ Eomes^+^ CD8 Tnn, another cell subset that can be seen in a state of T cell exhaustion(McLane et al., 2019). In both cases, these significant elevations in subset frequency are in a rare subset of cells but could represent the impact of decreased mucosal barrier function in IgA deficiency yielding increased and chronic antigenic stimulation via increased commensal translocation in the setting of reduced barrier function in SIgAD. This could be sufficient to yield altered T cell function, or consistent with recent findings(Lima-Junior et al., 2021), it is also possible that the inflammatory milieu caused by increased systemic accessibility of commensal microbes in the setting of fecal IgA deficiency leads to increased activity of endogenous retroviruses (ERV), which could in turn yield T cell exhaustion via chronic antigen stimulation in the setting of hyperinflammation. Recent evidence shows that chronic inflammation alone may be able to cause an exhausted-like phenotype (Wang, Nature Medicine 2019 and Porsche, JCI Insight, 2021). These hypotheses require future studies in our mouse model. Furthermore, detection of these shared signs of immune dysregulation in mouse and human reinforces the potential importance of our finding, though subtle, of T cell exhaustion-like changes in mouse and human. Of note, our findings in IgA deficiency are consistent with recent work showing evidence of T cell exhaustion in chronic inflammation(Maurice et al., 2021; Porsche et al., 2021; Wang et al., 2019). Of note, increased CD8 T cell expression of PD-1 was also reported in a recent pre-print using a mouse model of IgA deficiency (Gutzeit et al., 2021), demonstrating that this is not a single study phenomenon, though ours is the first evidence in human CD8 T cells. However, to fully test the hypothesis that IgA deficiency can increase the potential for the development of CD8 T cell exhaustion requires deeper investigation into T cell function, including the impact on antigen specific function, which remains to be clarified in future prospective studies with a larger cohort.

While IgA deficiency led to alterations in systemic immunity, the microbiome diversity and composition were indistinguishable between siblings who did and did not have IgA. Importantly, the lack of fecal microbiome dysbiosis in pediatric subjects with IgA deficiency suggests that systemic immune dysregulation in this population may be driven by aberrant exposures to commensal microbes rather than a normal response to a dysbiotic microbiome. In contrast, studies of adults report subtle microbiome differences between SIgAD and unrelated control subjects(Catanzaro et al., 2019; Fadlallah et al., 2018; Jorgensen et al., 2019; Moll et al., 2021). A strength of our study was using siblings living in the same household as controls to minimize environmental and genetic influences on the microbiome. These modest microbiome alterations in adult subjects with SIgAD could also reflect age-related changes that becomes evident later in life (e.g., changes in the microbiome secondary to disease-related chronic inflammation). There are limitations to our study which include the number of subjects (limited by recruitment of siblings with controls living in their home), potential for ascertainment bias and the possibility of species and strain level differences not resolved by 16s rRNA gene sequencing.

In this study, we developed an integrated approach (mFLOW-Seq) to simultaneously define the specific microbial targets of local secretory IgA and IgM and systemic IgG in children with IgA deficiency and their unaffected siblings, in addition to the host immunophenotype, investigating the impact of IgA deficiency on both cytokine milieu and lymphocyte subset presence and activation state. In our analysis of antibody targets, we adapted a Bayesian analytic approach(Jackson et al., 2021) to determine the probability of binding by each antibody isotype (i.e., IgA, IgM and IgG) to provide an intuitive and specific measure of antibody targeting across multiple isotypes. In comparison, previous studies used the ratio of the relative abundance of IgA+ and IgA-bacteria(Palm et al., 2014) or an IgA binding index(Kau et al., 2015) to define which microbes are targeted by IgA, each of which has drawbacks. The Bayesian approach provides several important advantages compared to other methods including avoiding artifactual associations based on different relative abundances of ASVs, increased power to detect differences in antibody binding to specific ASVs between experimental groups and lower coefficients of variation(Jackson et al., 2021). This study reveals clear connections at the level of common microbial targets across the mucosal (IgA and IgM) and systemic (IgG) antibody networks.

Sorted microbes into singly bound (i.e., IgA, IgM and IgG) and dually-targeted (e.g., IgA **and** IgM bound) groups revealed important properties of anti-commensal antibody networks using the same Bayesian strategy. First, we found that microbial populations from the same taxa exist in multiple binding states (e.g., unbound, singly bound by IgA, singly bound by IgM or dually bound by IgA and IgM). For example, *Coprococcus* has been described as a prominent IgA target in a previous study(Fadlallah et al., 2018), but we find a more complex pattern for this microbe with a 50% probability of being coated solely by IgM, 40% of being coated by IgM and IgA and less than 10% chance of being uncoated or coated by IgA alone. Second, the probability of IgA and IgM binding across the most abundant taxa follows a continuous distribution for most microbes with a few taxa (e.g., *Ruminococcus* and *Coprococcus)* being more highly IgA or IgM targeted, respectively. Almost all species of microbes are bound by secretory antibodies at least 25% of the time and few are bound more than 75% of the time. This may reflect cross-species reactivity(Pabst and Slack, 2020) or polyreactivity (Bunker et al., 2017) of many secretory antibodies to conserved epitopes like LPS and flagellin(Weis and Round, 2021). Third, microbes are dually coated by IgA and IgM at higher-than-expected proportions suggesting some degree of cooperative targeting of commensal microbes. Cerutti and colleagues also found dual IgA and IgM coating during their study of mucus associated microbes in adult IgA deficiency subjects (Magri et al., 2017). These findings suggest that once a microbe is coated by either IgA and/or IgM, it has an increased probability of being coated by a second immunoglobulin isotype. Sorting singly and dually bound microbes revealed a more complex biology that may help understand the relationship of mucosal IgA and IgM towards commensal microbes and how they collaborate in that process.

One of the remaining hurdles to achieve precision medicine is a mechanistic understanding of diseases where patients with a single “umbrella” diagnosis based on clinical findings or laboratory testing (i.e. IgA deficiency as currently defined based on serum testing) have diverse phenotypes, specifically in this case where only a small subset of patients are clinically symptomatic. Here, we use the disease of selective IgA deficiency to better understand how the gut humoral immune response tunes the baseline immunologic state. We hypothesize that the patients with more extreme immune dysregulation may be the patients with greater barrier dysfunction and they may in turn become symptomatic. However, while we think this study structure, based on paired affected and unaffected siblings, is ideal, being sufficiently powered to test this newly defined mechanistic hypothesis will require a multi-center study to identify and study sufficient families with the strategies used here. Here we have gained insight into the specific question of the impact of IgA deficiency on T cell function and cytokine environment, the broader question of how to study a disease where symptomatic patients are the exception rather than the norm and have provided a model with therapeutic implications to move this inquiry forward.

## Data Availability

Further information and request for resources and data should be directed to and will
be fulfilled by the lead contact, Michael Silverman.

## Acknowledgements

Thanks to the members of the Henrickson and Silverman labs for helpful discussions. We thank the staff at the mouse facility at Biomedical Research Building at the University of Pennsylvania. We thank the University of Pennsylvania Diabetes Research Center (DRC) for the use of the RIA Biomarker Core (P30-DK19525) and the CHOP Center for Human Phenomics (CHPS), supported by the National Center for Advancing Translational Sciences, National Institutes of Health (UL1TR001878). We thank the CHOP flow cytometry core for assistance with microbial flow cytometry and sorting and Penn-CHOP microbiome sequencing core for performing sequencing. We thank Dr. Ike Eisenlohr for helpful comments on the manuscript.

MAS, JS and SEH funded by CHOP Microbiome Pilot Grant. DAO was funded by NHLBI R38 HL143613 and NCI T32 CA009140. SEH funded by National Institute of Allergy and Infectious Diseases (NIAID) K08AI135091, Burroughs Wellcome Fund, the American Academy of Allergy, Asthma, and Immunology, the Immune Deficiency Foundation, the Primary Immune Deficiency Treatment Consortium and Chan Zuckerberg Initiative.

## Author Contributions

Author Contributions: Conceptualization, MAS and SEH; Methodology, PEC, KOB, LD, LV, KB, SEH, MAS; Investigation, PEC, LD, KOB, JG, JM, JBL, TD, CT; Writing – Original Draft, MAS and SEH.; Formal Analysis: PEC, LD, DO, SEH, MAS; Writing – Review & Editing, all authors; Visualization: MAS, LD, PEC.; Funding Acquisition, MAS, SEH and JS; Resources, DA; Supervision, MAS and SEH.

---

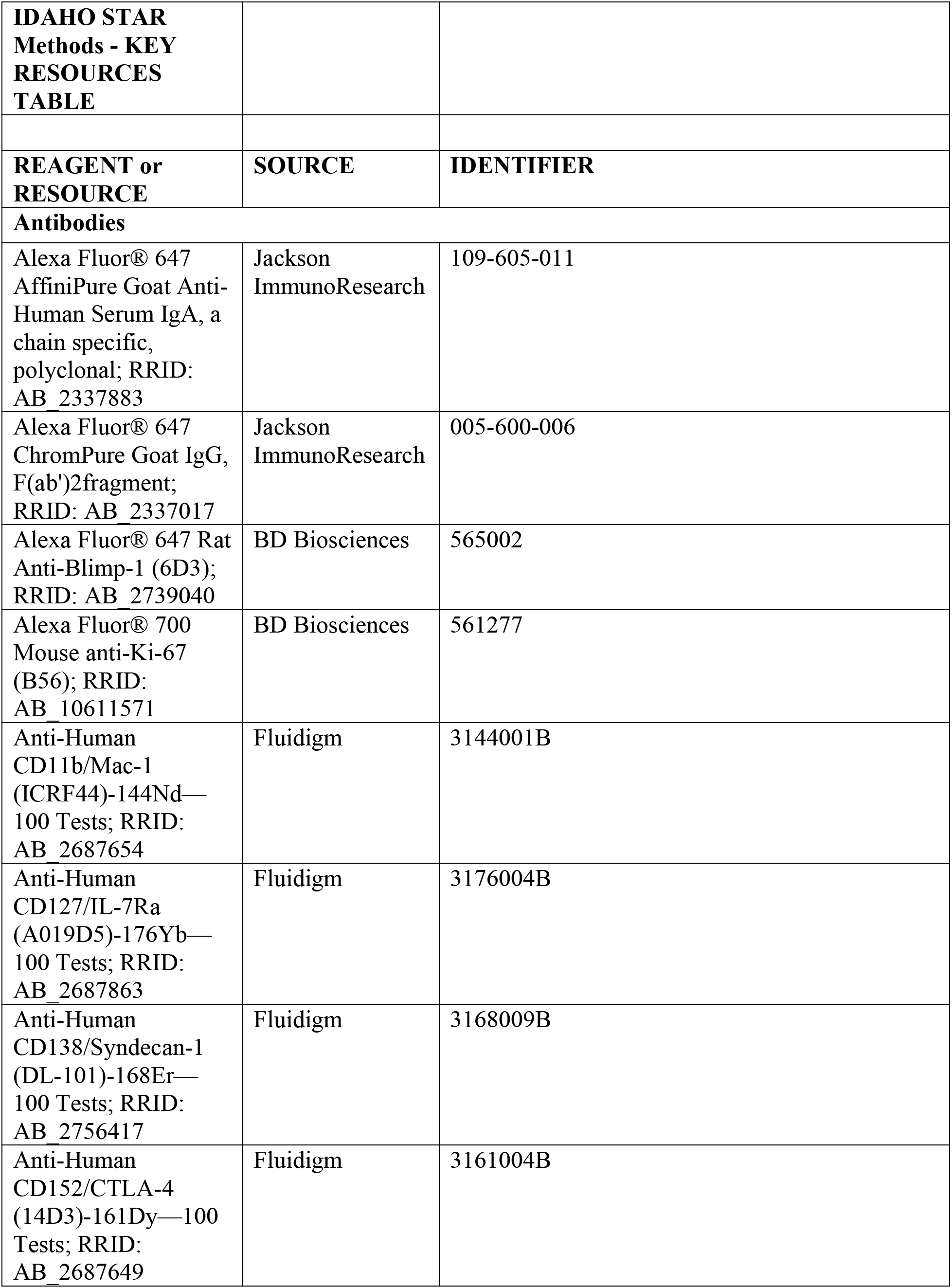
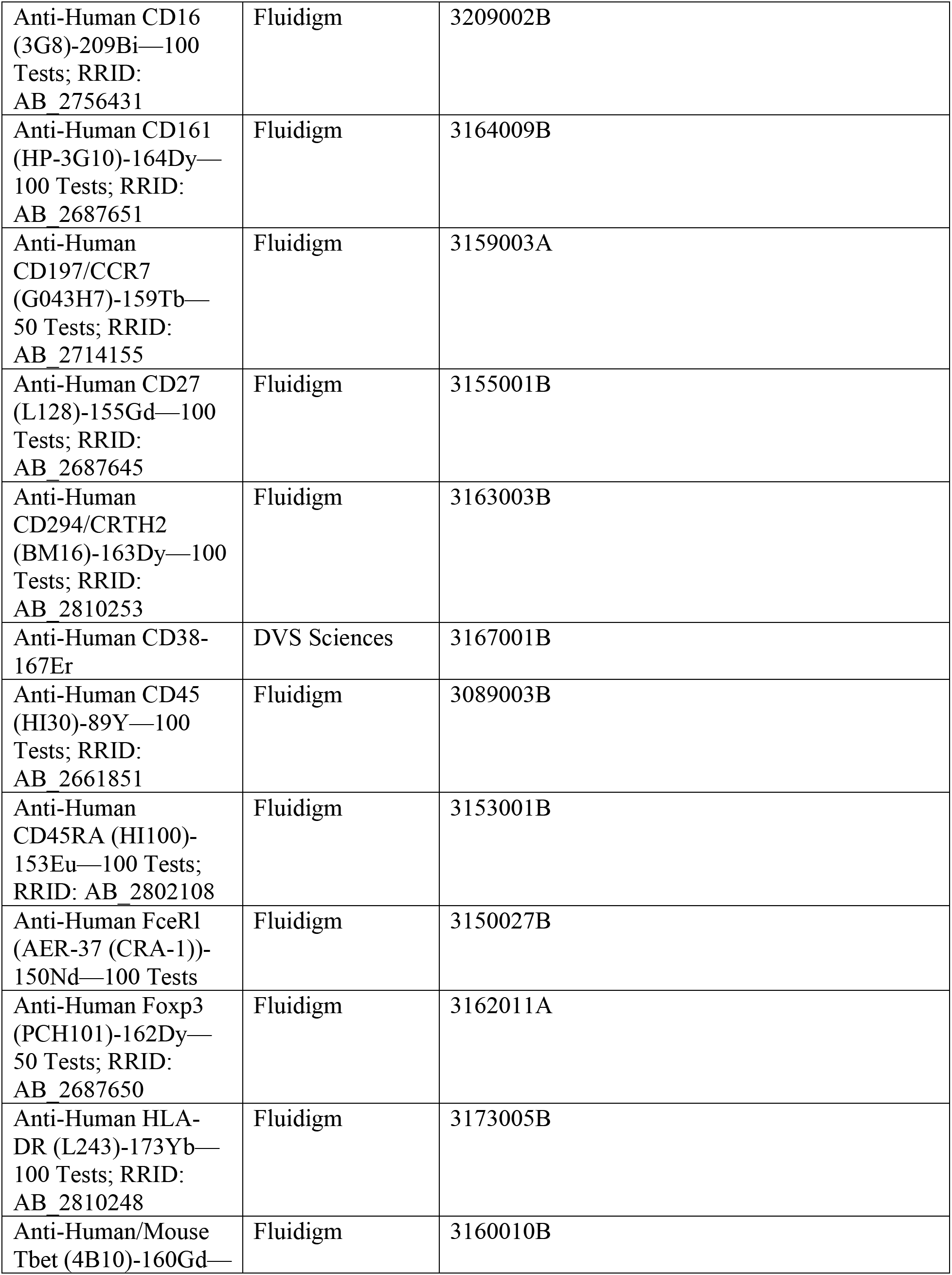
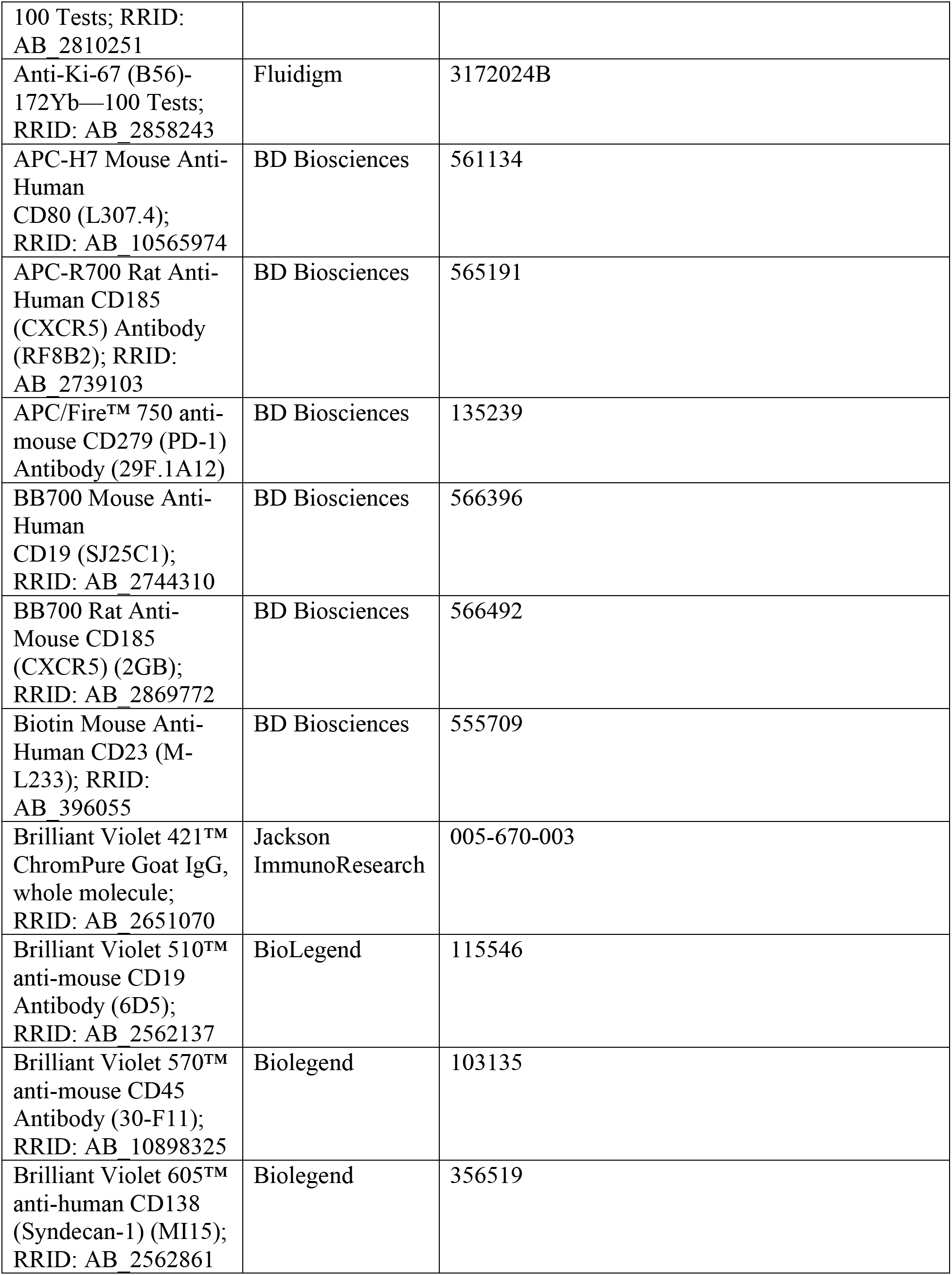
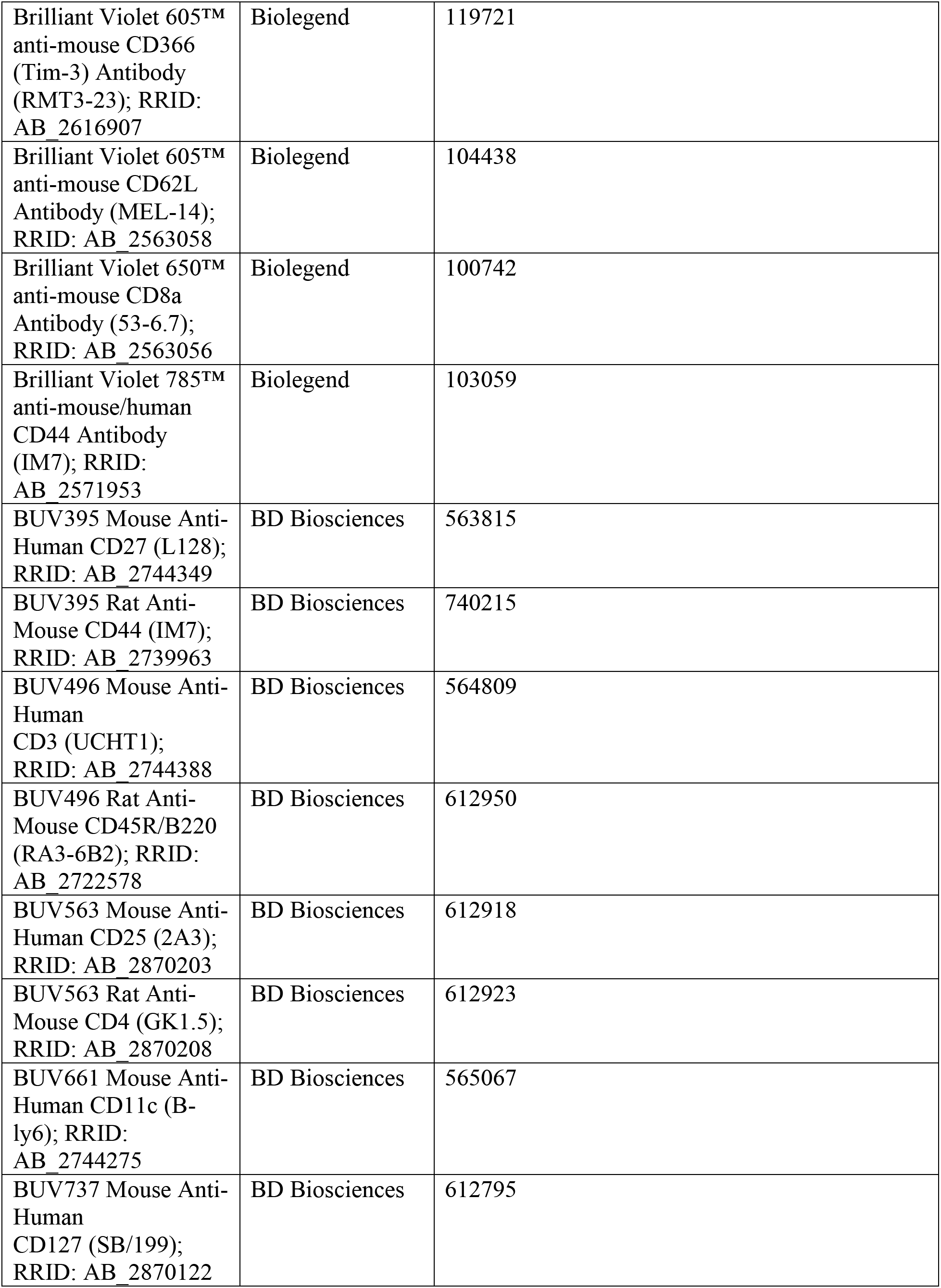
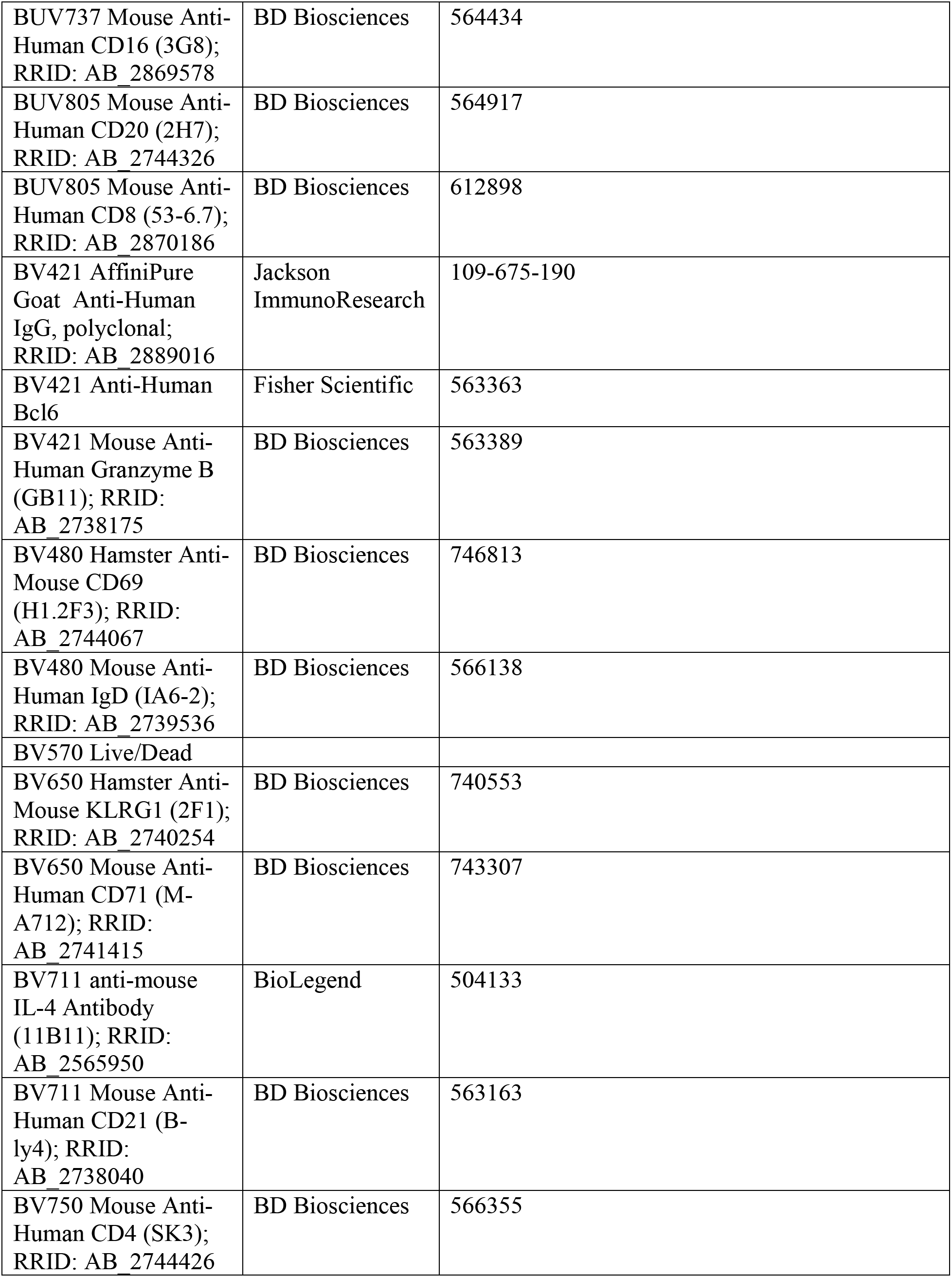
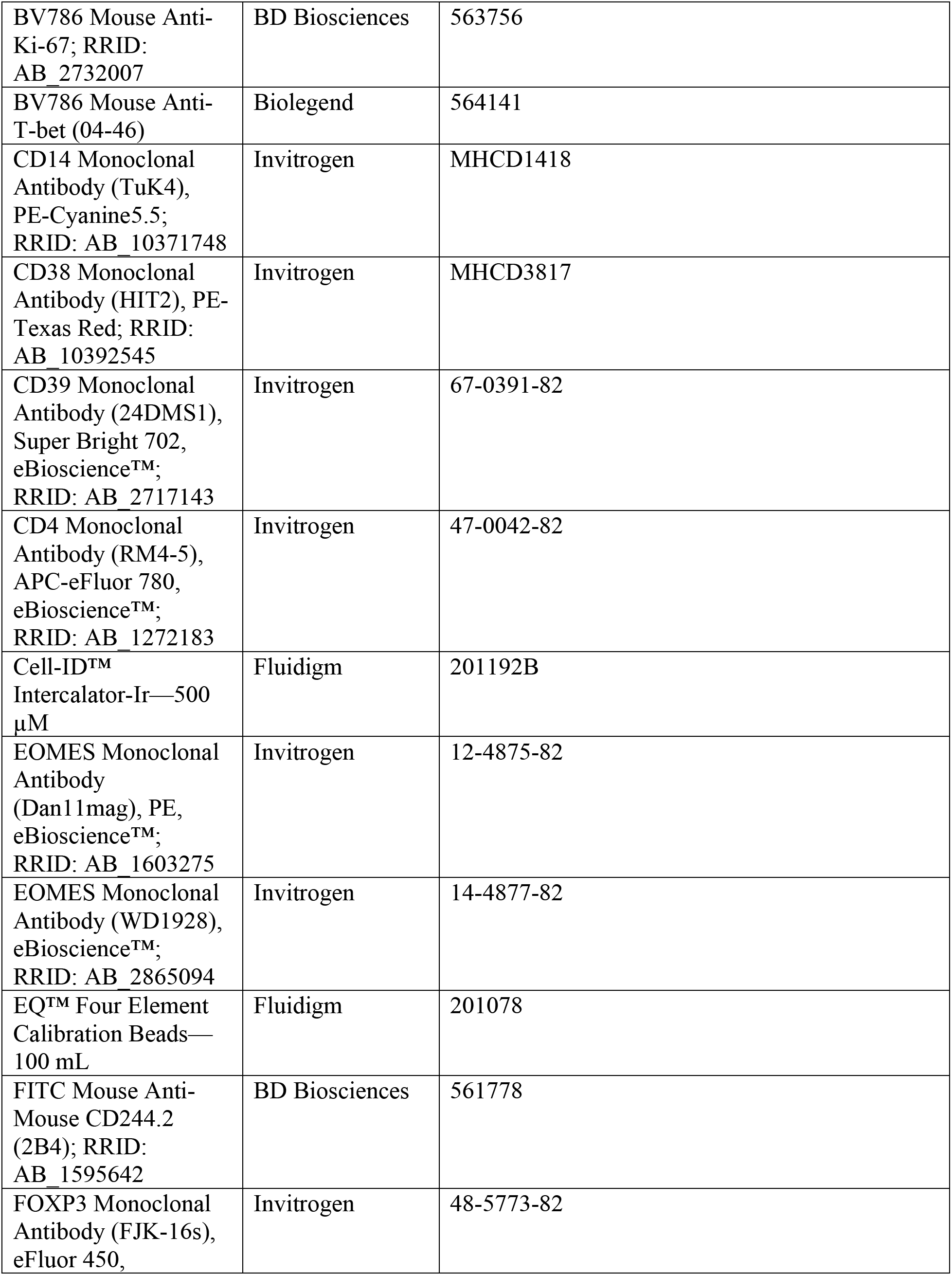
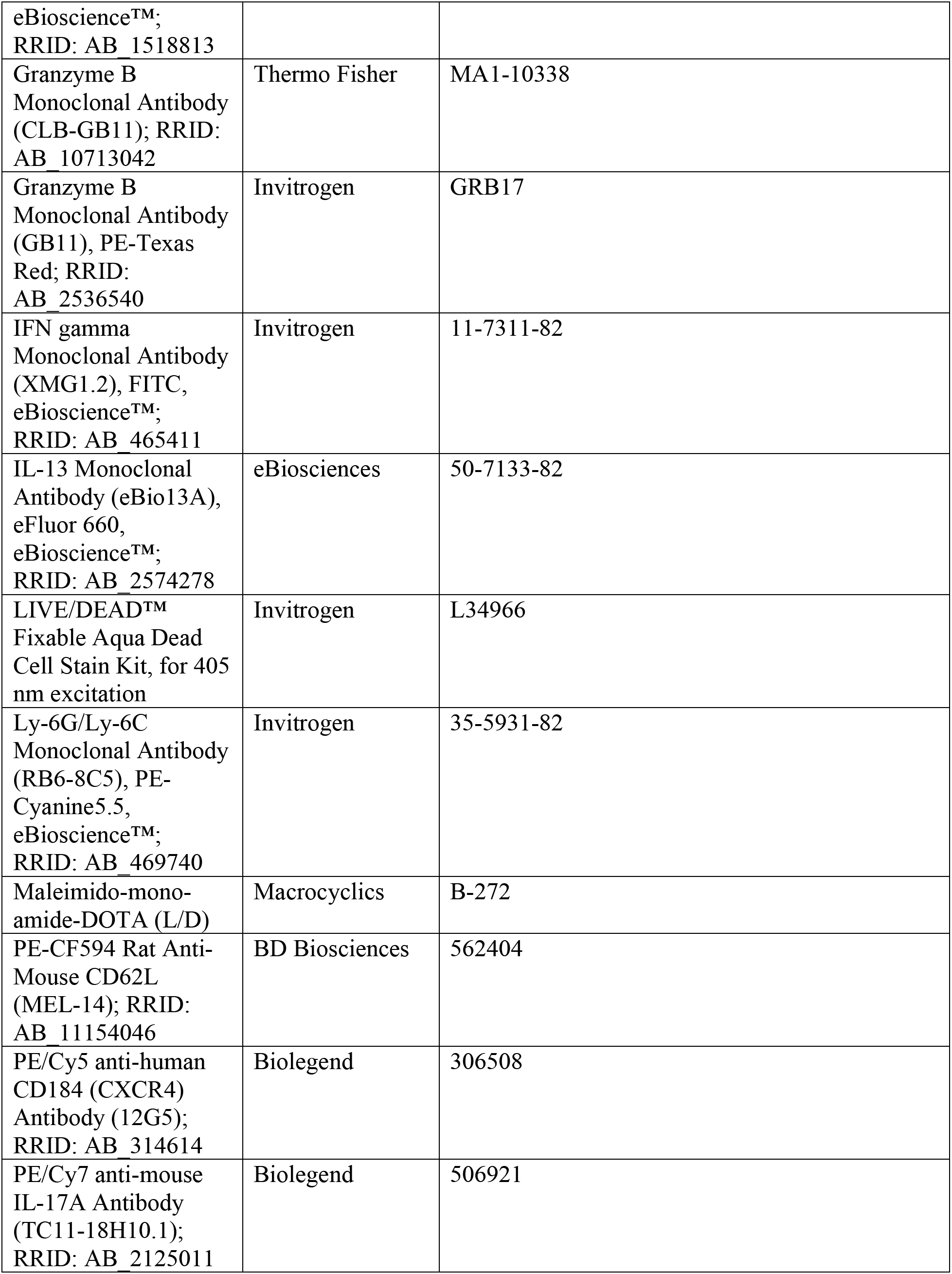
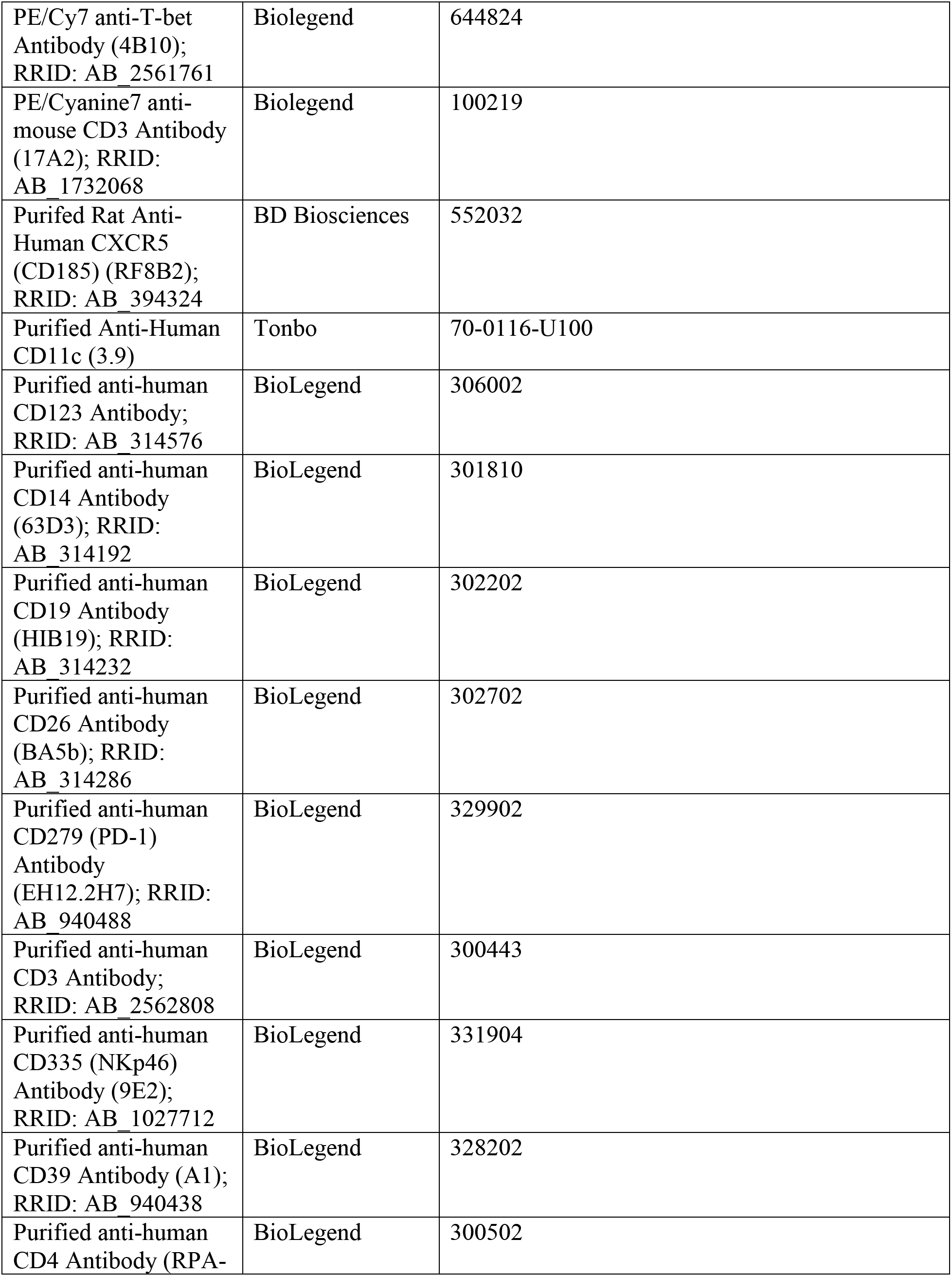
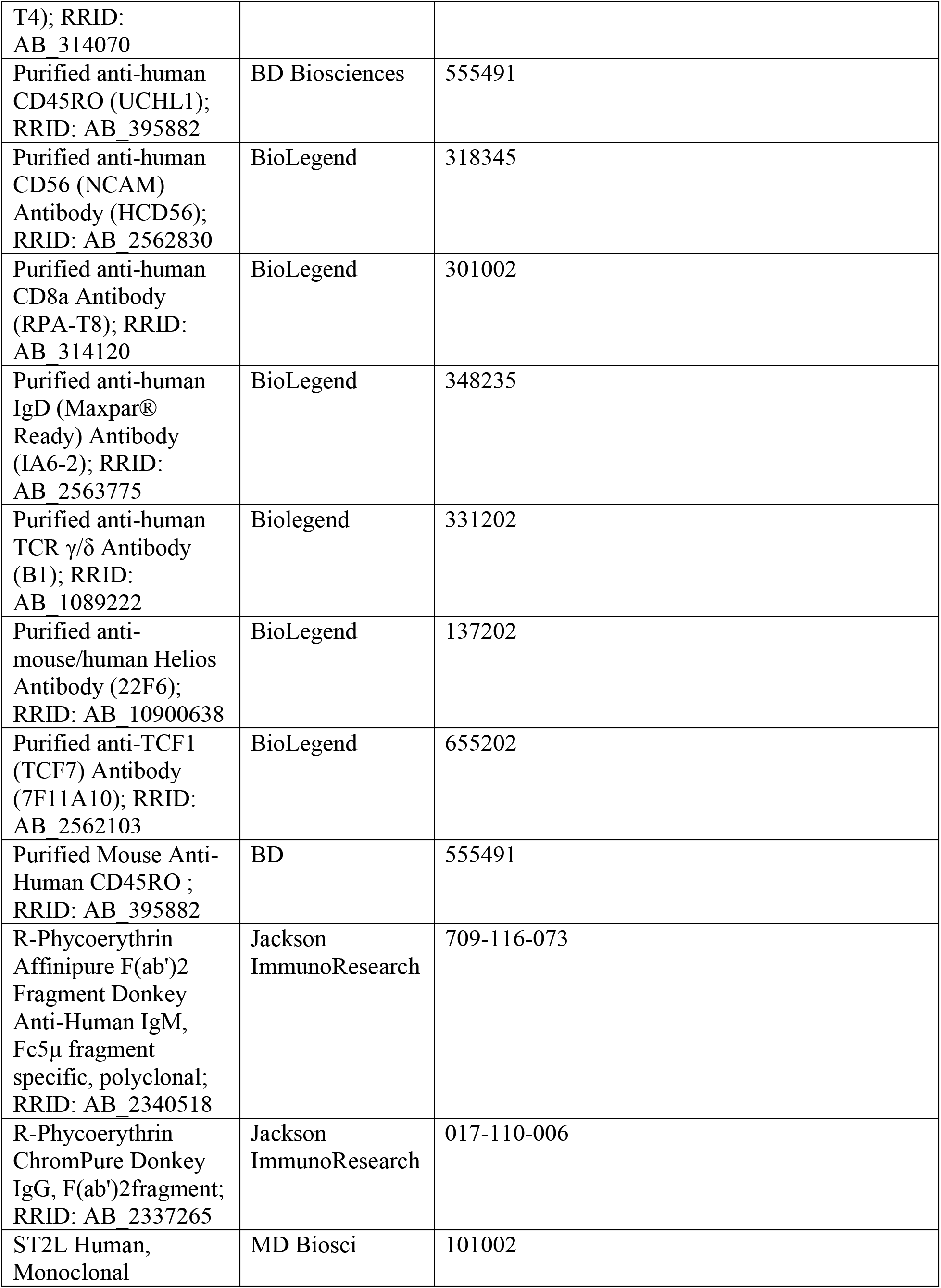
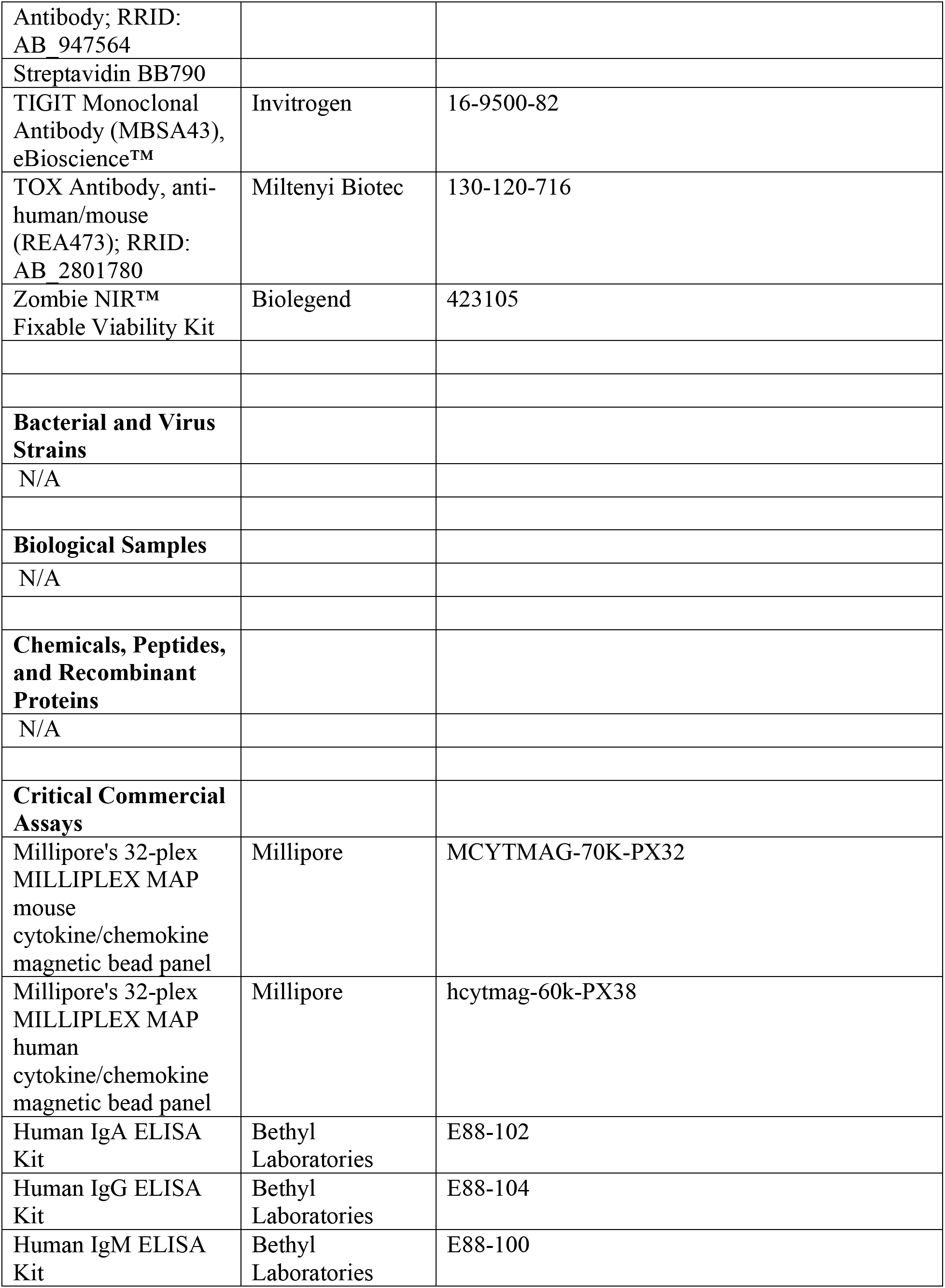
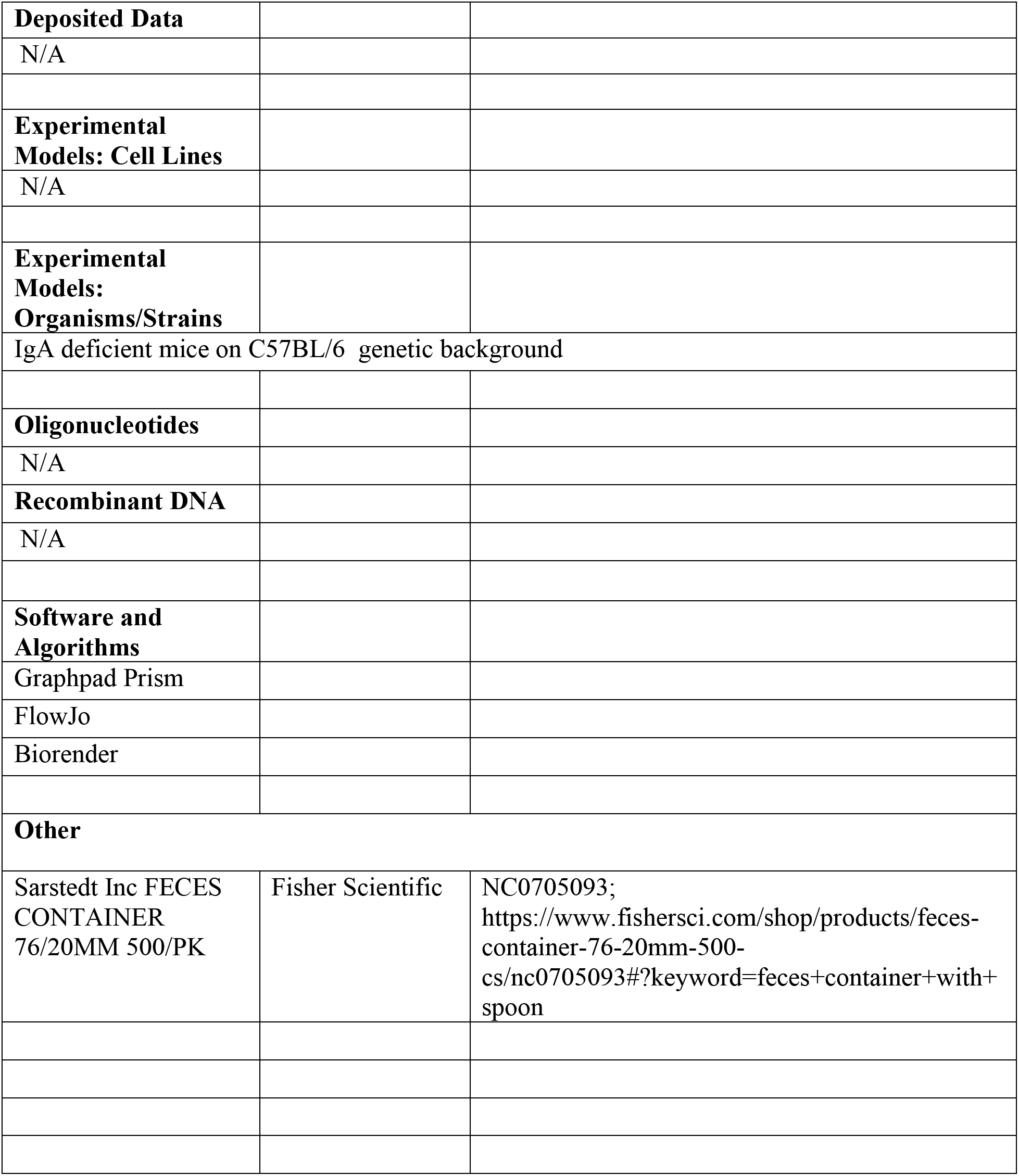

## STAR Experimental Methods

### Resource Availability

Further information and request for resources and reagents should be directed to and will be fulfilled by the lead contact, Michael Silverman

### Materials Availability

This study did not generate new or unique reagents.

### Data and Code Availability

All Code will be made publicly available using standard repositories such as GitHub.

#### i. Recruitment and sample collection

Patient recruitment began in the summer of 2017 and finished by the end of 2018. The Department of Biomedical Health Informatics (DBHI) and the Recruitment Enhancement Core (REC) assisted in developing a list of patients that were seen in the last 10 years by the Division of Immunology at The Children’s Hospital of Philadelphia with a diagnosis of IgA deficiency and a serum IgA level of <7 mg/dL. Families were contacted via email. For each family that was interested, a verbal screening consent was administered to allow for initial screening of patients by phone. We also reviewed the EMR of the IgA deficient sibling and non-deficient sibling to screen for inclusion and exclusion criteria. Once eligibility was confirmed, the family was scheduled for an initial study visit. In the spring of 2018, a final effort was made to recruit any newly diagnosed patients using the same criteria. Letters were then sent via post mail to any families from the initial list that were not able to be contacted via email or phone and any newly identified patients from the final list. If we did not receive a response within 2 weeks after the letters were sent, we followed up with the family via telephone. The same process was used for verbal consent and patient screening for inclusion and exclusion criteria. A total of 220 emails and letters were sent during the recruitment period (communications comprised of 3 email blasts and 1 round of letters). Initial participant IDs that were given to the consented families were recoded and only known to the study team.

#### ii. PBMC sample processing

Peripheral blood mononuclear cells (PBMCs) were isolated using Ficoll density gradient. Whole blood was mixed 1:1 with PBS and layered onto Ficoll gradient. Cells were centrifuged. Ficolled plasma was saved for later analysis. Buffy coat suspension was removed and spun down for cell isolation. ACK lysis was performed. Cells were counted in RPMI (10% FBS, 1% L-Glutamine, 1% Pen-Strep). Cells were spun down and resuspended in 1 mL of freezing media (90% FBS, 10% DMSO).

#### iii. Bacterial flow cytometry

Stool pellets were homogenized in 1000µL of sterile-filtered PBS with 1% bovine serum albumin (BSA) (Fisher). The homogenates were centrifuged at 100 x g for 15 minutes. The supernatants were transferred to sterile 1.5-mL Eppendorf tubes and centrifuged at 8000 x g for 5 minutes. Bacterial pellets were resuspended in 1000µL of PBS with 1% BSA and measured for optical density (OD600) with a spectrophotometer (BioPhotometer, Eppendorf). All samples were normalized to 0.1 OD by addition of appropriate volume of PBS with 1% BSA. Bacteria were fixed in 500µL of 4% paraformaldehyde (PFA) solution for 20 minutes at room temperature. Bacteria were then washed and transferred to a 96-well U-bottom plate (CellTreat) and incubated in blocking solution (PBS, 1% BSA and 20% normal rat serum) for 30 minutes at 4C. Serum diluted in bacterial staining buffer (BSB) (PBS, 1% BSA, and 0.05% Sodium Azide (VWR)) to a final IgG concentration of 10µg/mL were added to the fixed bacteria and incubated overnight. Bacteria were washed once in BSB and staining was performed with fluorochrome-conjugated polyclonal antibodies: IgA-AF647, IgG-BV421, and IgM-PE (Jackson Immuno Research), diluted 1:100, 1:200, and 1:400 respectively in BSB, for 20 minutes at 4C. Bacteria were washed in Tris Buffered Saline (TBS) with 1% BSA and incubated in SYTO BC green fluorescent nucleic acid stain (Thermo Fisher), diluted 1:500 in TBS, for 15 minutes at room temperature. Bacteria were washed in BSB and transferred to FACS tubes (Falcon). Flow cytometric analysis was performed on the LSRFortessa using the standard excitation lasers (violet-405, blue-488, yellow-green-561, and red-640) and emission filters (violet-450/50, violet-710/50, blue-525/50, yellow-green-582/15, yellow-green-780/60, and red-660/20). The FSC and SSC thresholds were set at 200. To correctly set the voltages and identify the appropriate size gates, unstained fecal microbes and sterile-filtered BSB were used as controls in each experiment. Microbes were initially gated by FSC-A and SSC-A, then selected for SYTO BC positive events. For each sample, isotype stained controls were used to define the gates for IgA, IgM and IgG coated microbes.

#### iv. Sibling swap flow cytometry

Bacteria were isolated from stool pellets as described in iii. After the blocking step, bacteria were incubated with subject’s own serum or sibling’s serum. Staining and flow cytometric analysis was performed as described in iii.

#### v. Flow cytometric cell sorting

Fecal microbes were isolated from stool pellets and processed as described above in iii. Populations of interest were identified and gated as described above in iii on BD FACSAria II and double sorted into 1.5 mL Eppendorf tubes (100,000 cells per sample). First, sample buffer without fecal bacteria was run to establish a background. Second, an unstained fecal bacterial sample was run to identify bacteria by physical parameters (FSC and SSC) and to set initial negative gates. Third, a sample with Syto BC green fluorescent nucleic acid stain (Thermo Fisher) was run to identify bacteria gate. For each subject, an isotype stained fecal sample was used to fine tune the negative gates. Fourth, the patient sample with stool staining (no serum) was run and sorted as described in iii. Immediately following, the patient sample with stool and patient serum was run and sorted. Lastly, purity checks were run on most populations to validate each sort.

#### vi. Fecal ELISA

Fecal microbes were isolated from fecal pellets as described above with slight modification. After the second centrifugation step, the supernatant was used to measure the concentration of free immunoglobulin in the gut while the pellets were used to measure the bacterial-bound immunoglobulin concentration. The assay was performed using the Human ELISA Quantitation Set according to manufacturer instructions (E80-102, E80-104, E80-100; Bethyl).

#### vii. 16S rRNA gene sequencing and analysis

The 16S rRNA gene sequencing of the V4 variable region was performed at the Children’s Hospital of Philadelphia sequencing core on the Illumina MiSeq platform using the protocol previously described [2]. The data was processed using QIIME[v2018.2], denoised and clustered into Amplicon Sequence Variants (ASVs) using DADA2. Taxonomic classification was generated using Greengenes 99% reference database v13.8. To determine 1-diversity, weighted and unweighted UniFrac analysis were performed between each sample time points and sample sources. Differences in alpha diversity between study groups or populations were calculated using linear mixed effects models using the subject IDs as random effects. Beta diversity was visualized using principal coordinate plots. Difference in centroids between groups were calculated using PERMANOVA test. The binding probability ratios of each ASV was calculated for the IgA, IgM and IgG populations using the method described before(Jackson et al., 2021) with the following modifications. Differences between binding probability ratios were estimated using linear mixed effects models. Individual probabilities were obtained using ALR transformations on the probability ratios. The total binding probability of an antibody isotype was then defined as the sum of double positive probability and single positive probability. Binding probabilities were plotted for antibody isotype pairs for each ASV. Preferential binding was then determined by calculating the angle the data point makes with the x axis and determining if it is significantly higher or lower than 45° using one tailed t-test. Multiple tests were corrected for false discovery rate using Benjamini-Hochberg method.

#### viii. Statistical analysis

Data are presented as medians. Statistical significance was determined as indicated in the figure legends with Prism 9 (GraphPad Software Inc). P values <0.05 were considered statistically significant. Linear regressions, Pearson, Spearman correlations were calculated using Prism 9. Heatmaps were generated using publicly available software from the Broad Institute (Morpheus, https://software.broadinstitute.org/morpheus). Analytes with less than 50% of values above the limit of detection in both cases and control groups were excluded from analysis. For outlier analysis, Z-scores were first computed for each analyte (across patients), and the corresponding Z-score vectors (across analytes) were then transformed to a patient-level chi-square statistic computed by the formula of Fisher’s method of metanalysis using a standard normal reference distribution and two-tailed assumptions. P-values for outlier significance were computed by permutation test, whereby the patient label was permuted separately for each individual analyte to create a reference distribution for the resulting chi-squared statistic.

#### ix. Mass cytometry sample staining and acquisition

Cryopreserved PBMCs were thawed in 10mL cRPMI (RPMI 1640 1X supplemented with 10% heat-inactivated fetal bovine serum (FBS), 1% L-glutamine and 1% Pen-Strep). PBMCs were centrifuged at 315 xg for 5 min and resuspended at a concentration of 5 x 106 cells/mL in complete medium. Cell suspensions were plated at 200uL per well in a 96-well flat-bottom plate and incubated for 1 h at 37°C to let cells rest. After 1-hour incubation, cell suspensions were transferred to a 96-well round-bottom plate. Plate was centrifuged at 515 xg for 5 min. Cells were resuspended in 50uL of live/dead mDOTA-La139 (Macrocyclics, stock 41.5mg/mL, used at 1:4000) and incubated at room temperature for 5-10 minutes. Cells were washed with stain buffer (PBS, 1% FBS) and centrifuged at 515 xg for 5 min. Surface proteins were stained by resuspending cells in 50uL of surface antibody cocktail and incubated at room temperature for 30 mins. Cells were washed a further two times with stain buffer and centrifuged at 515 xg for 5 min. Cells were permeabilized with 50uL of a permeabilization reagent which included 3 parts Fixation/Permeabilization Concentrate (Invitrogen) and 1 part eBioscience Fixation/Perm Diluent (Invitrogen). After 30 minutes of incubation, cells were washed with 1x PERM buffer (1 part 40X Permeabilization concentrate, 9 parts H2O) and centrifuged at 650 xg for 5 min. Intracellular proteins were stained by resuspending cells in 50uL of intracellular antibody cocktail and incubated at room temperature for 1 h. Cells were washed three times with 1x permeabilization buffer and centrifuged at 650xg for 5 min. Cells were fixed with 100uL of 1.6% PFA (Alfa Aesar, Fisher Scientific) and 125nM Iridium (Fluidigm, used at 1:4000) and stored overnight at 4°C. Prior to acquisition, cells were washed with twice with PBS and twice with Milli-Q H2O.

#### x. Flow cytometry sample staining and acquisition

Cryopreserved PBMCs were thawed in 10mL cRPMI (RPMI 1640 1X supplemented with 10% heat-inactivated fetal bovine serum (FBS), 1% L-glutamine and 1% Pen-Strep). PBMCs were centrifuged at 315 x g for 5 minutes and resuspended at a concentration of 2 x 10^6^ cells/mL in complete medium. Cell suspensions were plated at 200uL per well in 96-well round-bottom plates and centrifuged at 515 x g for 5 minutes. Cell pellets were resuspended in 50uL of surface antibody cocktail and incubated at room temperature, in the dark, for 20 minutes. Cells were washed with 150uL FACS buffer (PBS, 1% FBS) and spun down at 500 x g for 5 minutes. Cells were permeabilized with 50uL of a permeabilization reagent which included 3 parts Fixation/Permeabilization Concentrate (Invitrogen) and 1 part eBioscience fixation/perm diluent (Invitrogen). Cell suspensions incubated for 20 minutes at room temperature followed by a wash with 150ul PERM Buffer (1 part 10X permeabilization concentrate and 9 parts water). Cell suspensions were centrifuged at 500 x g for 5 minutes and resuspended in 50uL of intracellular antibody cocktail. Cells incubated for 60 minutes at room temperature and were subsequently washed with 150uL of permeabilization buffer. Cells were resuspended in 100uL of a streptavidin-BB 790 antibody cocktail and incubated for 10 minutes before fixing with 1.6% PFA. Fixed cells were stored overnight at 4°C and analyzed on a BD LSR, Fortessa or X-50 the following day.

#### xi. Fecal DNA extraction

Bacterial DNA was isolated from fecal samples using the DNAeasy PowerSoil HTP 96 kit according to manufacturer instructions (12955-4; Qiagen)

#### xii. Low biomass DNA extraction

Sorted samples underwent three freeze-thaw cycles followed by a 2X SDS Lysozyme Proteinase-K (SLP) lysis buffer (50 mM Tris, 50 mM EDTA, 50 mM sucrose, 100 mM NaCl) treatment. Samples were then treated with 0.2 volume 50mg/mL of lysozyme and incubated at 37C for 30 minutes. Following lysozyme treatment, 0.2 volume Proteinase K (20mg/mL) and 0.1 volume 10% SDS was then added, and samples were incubated overnight at 56C. On Day 2, DNA was isolated with 5 M NaCl and isopropanol precipitation with 40 *μ*g of glycogen as a DNA carrier. Isolated DNA was stored at −20C prior to analysis.

#### xiii. Human and murine serum cytokine analysis

Millipore’s 38-plex MILLIPLEX MAP human cytokine/chemokine magnetic bead panel kit (hcytmag-60k-PX38; Millipore) was used to measure the following cytokines and chemokines: IL-1a, IL-11, IL-1Ra, IL-2, IL-3, IL-4, IL-5, IL-6, IL-7, IL-8, IL-9, IL-10, IL-12 (p40), IL-12 (p70), IL-13, IL-15, IL-17, IP-10, EGF, eotaxin, FGF-2, Flt-3 ligand, fractalkine, G-CSF, GM-CSF, GRO, IFN-a2, IFN-y, MCP-1, MCP-3, MDC, MIP-1-a, MIP-11, sCD40L, TGFa, TNFa, TNF1, and VEGF. The immunoassay procedure was performed according to the manufacturer’s instructions and the plate was run on a MAGPIX multiplex reader which was analyzed with Luminex xPONENT software. Concentrations are given in picograms per milliliter (pg/mL). Millipore’s 32-plex MILLIPLEX MAP mouse cytokine/chemokine magnetic bead panel kit (MCYTMAG-70K-PX32; Millipore) was used to measure the following cytokines and chemokines: G-CSF, GM-CSF, IFN-y, IL-1a, IL-11, IL-2, IL-3, IL-4, IL-5, IL-6, IL-7, IL-8, IL-9, IL-10, IL-12 (p40), IL-12 (p70), IL-13, IL-15, IL-17, IP-10, KC, MCP-1, MIP-11, M-CSF, MIP-2, MIG, RANTES, VEGF and TNFa. The immunoassay procedure was performed according to the manufacturer’s instructions and the plate was run on a MAGPIX multiplex reader which was analyzed with Luminex xPONENT software. Concentrations are given in picograms per milliliter (pg/mL). Limit of detection is 3.2 pg/mL. Values that were listed as <3.2 where the actual read net fluorescence is lower than the lowest standard net reading were set to 3.2 pg/ml.

#### xiii. Animal studies

IgA^-/-^ and IgA^+/-^ mice were generated by breeding IgA^-/-^ mice to congenic C57BL/6 mice born and maintained in the BRB facility at the University of Pennsylvania (provided by Dr. Allman(Wilmore et al., 2018)). IgA^+/-^ sires were then mated to IgA^-/-^ dams to generate experimental mice which include littermates of both sexes age 5-14 weeks. We utilized this breeding scheme to standardize maternal antibody and microbiome transfer effects and to avoid cage effects(Rogier et al., 2014). All experiments were approved by IACUC. Spleens, mesenteric lymph nodes (MLN) and serum was collected from IgA deficient and heterozygous mice derived from breeding of IgA deficient dam to IgA heterozygous sire in most cases. Spleens and MLNs collected from mice were smashed onto 70*μ*m filters, rinsed with RPMI + 10% FBS medium (RPMI 1640 supplemented with 10% heat-inactivated fetal bovine serum (FBS)) and centrifuged at 400xg for 4 mins at 4°C. Splenocytes were treated with ACK lysis buffer (Gibco) for 90 seconds. Cells were centrifuged at 400xg for 4 mins at 4°C and washed with RPMI +10% FBS and counted on Countess cell counter with Trypan blue stain (0.4%). For chemokine panel staining, splenocytes were plated in a round bottom 96-well plate at 2 x 10^6^ splenocytes/well. After centrifugation, cells were stained with Zombie-NIR viability dye for 15 mins at RT. Cells were washed once with FACS buffer and resuspended in surface stain fluorescent antibody cocktail mix and incubated at RT. After 1hr, cells were washed with FACS buffer, fixed and permeabilized for 30 mins at 4°C with BD Cytofix/Cytoperm kit (BD Biosciences). After 30 mins, cells were washed with FACS buffer and resuspended in intracellular stain fluorescent antibody cocktail mix and incubated for 1hr at 4°C. Cells were fixed in 1.6% PFA overnight and analyzed on a 5-laser Cytek Aurora after 24hrs. For cytokine panel staining, splenocytes were plated in a flat bottom 96-well plate at 2 x 10^6^ splenocytes/well and stimulated for 4 hours at 37°C in the presence of PMA/Ionomycin as well as BFA (GolgiPlug; BD Biosciences) and Monensin (GogliStop; BD Biosciences). After 4hrs, cells were washed with FACS buffer, centrifuged at 400xg for 4 mins at 4°C and resuspended in surface stain fluorescent antibody cocktail mix with the addition of Live/Dead Aqua viability dye. After 30 mins, cells were washed with FACS buffer, fixed and permeabilized for 30 mins at 4°C with BD Cytofix/Cytoperm kit (BD Biosciences). After 30 mins, cells were washed with FACS buffer and resuspended in intracellular stain fluorescent antibody cocktail mix and incubated for 1hr at 4°C. Cells were fixed in 1.6% PFA overnight and analyzed on a 5-laser Cytek Aurora after 24hrs. For serum cytokine/chemokine studies, serum was obtained from the retro-orbital venous plexus at the time of sacrifice and then frozen at −80°C.

#### xiv. Vaccine Assays

Blood was collected into sodium heparin tubes as above and processed by CHOP Translational Core Laboratory for PBMCs and plasma. Research testing on subject plasma for diphtheria and tetanus antibody titers was performed by CHOP central lab. Subject plasma was sent to ARUP Laboratories in Salt Lake City, UT to perform research measurements of Measles IgG, Mumps IgG and *S. Pneumoniae* titers.

