## Supplemental Files for "IgA deficiency destabilizes immunological homeostasis towards intestinal microbiota and increases the risk of systemic immune dysregulation"

### Supplemental Figure Legends

**Figure S1. Secretory IgA and IgM and systemic IgG coat diverse microbes in healthy children, and overlapping sets of microbes are targeted in healthy and fecal IgA deficient children; Related to Figure 1. A.** Percentage of fecal microbes bound by IgM and IgA in healthy participants. **B.** Microbial flow cytometry (mFLOW) schematic gating and sorting scheme. Fecal microbes were incubated with autologous serum (normalized to 10 ug/mL of IgG) stained with the nucleic acid dye Syto BC and either fluorophore conjugated anti-IgA, anti-IgM or anti-IgG antibodies or isotype controls. Each population was sorted by FACS. 100,000 cells from each population were sorted twice and then checked for purity. Sibling pairs were sorted on the same day using the same ARIA sorter. DNA from 14 sorted population were then extracted and microbiome amplification, sequencing and analysis were completed**. C.** Shannon alpha diversity and **D.** Principal coordinates analysis (PCoA) of unweighted UniFrac distances of unsorted fecal microbiomes from control and fecal IgA deficient siblings. **E.** Taxa bar plots of the relative abundances of fecal microbes at the family taxonomic level. **F-G.** Alpha diversities of the microbiomes from the indicated sorted populations.

**Figure S2. Secretory IgA and IgM target an overlapping set of microbes in healthy children, Related to Figure 1. A-B.** Plots showing ASVs targeted by IgA and IgM. **C.** ASVs dually targeted by IgA and IgM. **D.** Venn diagrams illustrating the relationship of IgM+IgA-, IgM+IgA+ and IgM-IgA+ positive sorted populations of microbes from control siblings. Linear mixed effect (LME) modeling identified microbial taxa differentially coated by IgA and/or IgM in each sorted fraction (FDR<0.05).

**Figure S3. Systemic IgM and IgG target an overlapping set of microbes in healthy children; Related to Figure 1. A-B.** Plots showing ASVs targeted by IgG and IgM. **C.** ASVs dually targeted by IgG and IgM. **D.** Venn diagrams illustrating the relationship of IgM+IgG-, IgM+IgG+ and IgM-IgG+ positive sorted populations of microbes. Linear mixed effect (LME) modeling identified microbial taxa differentially coated by IgG and/or IgM in each sorted fraction (FDR<0.05).

**Figure S4. Serum IgA concentration does not correlate with fecal IgA concentration or the percent of bacteria bound by IgA; Related to Figure 3. A.** ELISA of serum IgA concentration of control (n=13) and SIgAD participants (n=19). **B.** ELISA of free and bacteria-bound secretory IgA concentration from fecal samples. **C.** Microbial flow cytometry plots of fecal microbes stained with anti-IgA and anti-IgM fluorophore conjugated secondary antibodies from two pairs of control and SIgAD siblings. **D.** Percentage of fecal microbes coated with IgA. **E.** Flow cytometry of human PBMCs, stained for surface IgA and IgG on memory B cells (CD19+ CD27+). **F.** Cartoon illustrating fecal and serum IgA status of the 32 subjects in this cohort. **G.** Correlation of the percentage of IgA coated microbes and serum IgA concentration (left panel); free IgA concentration in fecal water and serum IgA (middle panel), and bacteria bound IgA and serum IgA concentration (right panel). Spearman correlation, line represents simple linear regression. *P<0.05, **P<0.01, ***P<0.001, ****P<0.0001.

**Figure S5. Serum IgG binds to a higher proportion of microbes in fecal IgA deficient humans and mice compared to controls; Related to Figure 4. A.** Serum IgM concentration of fecal IgA+ and fecal IgA- study participants. **B.** Percentage of circulating IgM+ memory B cells (CD19+ CD27+). **C.** Percentage of fecal microbes coated by IgM or IgG between fecal IgA+ and fecal IgA- participants. **D**. Mean fluorescent intensity (MFI) of IgG bound to stool from fecal IgA+ and IgA- participant stool. **E.** Percentage of IgG binding to microbes in mFLOW is shown for three sibling pairs. Serum IgG concentration was determined by ELISA and then serially diluted to concentrations 0.001 to 10,000 ug/mL. Diluted serum was then incubated with fecal microbes, stained with fluorophore conjugated anti-IgG secondary antibodies and analyzed by microbial flow cytometry to determine the percentage of microbes bound by each antibody isotype. Each color line represents a family. Circles are fecal IgA deficient probands and squares are healthy control siblings. **F.** Serum and fecal microbiota from IgA deficient and heterozygous littermates were combined. Autologous combinations (serum and feces from IgA KO or serum and feces from IgA heterozygous mice) or non-autologous (serum from IgA KO and fecal microbes from IgA heterozygous mouse, or serum from heterozygous mouse and fecal microbes from IgA KO mouse). K=KO and H=heterozygous littermate. “K-H” indicates IgA KO fecal microbes incubated with IgA heterozygous serum. “H-K” indicates IgA heterozygote fecal microbes incubated with IgA KO serum. *P<0.05

**Figure S6. Cellular immune-phenotyping in IgA deficient humans and mice; Related to Figure 6. A.** Peripheral blood CD4 T helper subsets analyzed by CyTOF in fecal IgA+ and fecal IgA- participants. **B**. Heatmap of exhaustion-like clusters from PBMCs of human fecal IgA- and IgA+ participants. **C**. Tbet expression on non-naïve CD8 T cells from mesenteric lymph nodes in $IgA^{-/-}$ mice versus $IgA^{+/-}$ control mice. **D.** Splenic T cell cytokine expression from $IgA^{-/-}$ mice versus $IgA^{+/-}$ control mice after stimulation with PMA/Ionomycin. **E.** Percent positivity for measles and mumps titers. Diphtheria and tetanus antibody levels. Percent positivity of *S. pneumoniae* vaccine titers.

**Figure S7. Fecal IgA deficient humans and mice display serum cytokine and chemokine dysregulation; Related to Figure 7. A.** Serum cytokine concentrations in patients with fecal IgA deficiency (n=15) and their healthy siblings (n=17). **B.** Serum cytokine concentrations between $IgA^{-/-}$ mice versus $IgA^{+/-}$ mice. **C.** QQ plot illustrating observed versus expected chi-squared statistics computed by Fisher’s method of meta-

analysis across cytokines for patient outlier detection. Analysis includes all 38 cytokines.
**D.** Levels of serum anti-LPS antibodies measured by ELISA.

**Figure S1. Secretory IgA and IgM and systemic IgG coat diverse microbes in healthy children, and overlapping sets of microbes are targeted in healthy and fecal IgA deficient children, related to Figure 1.**

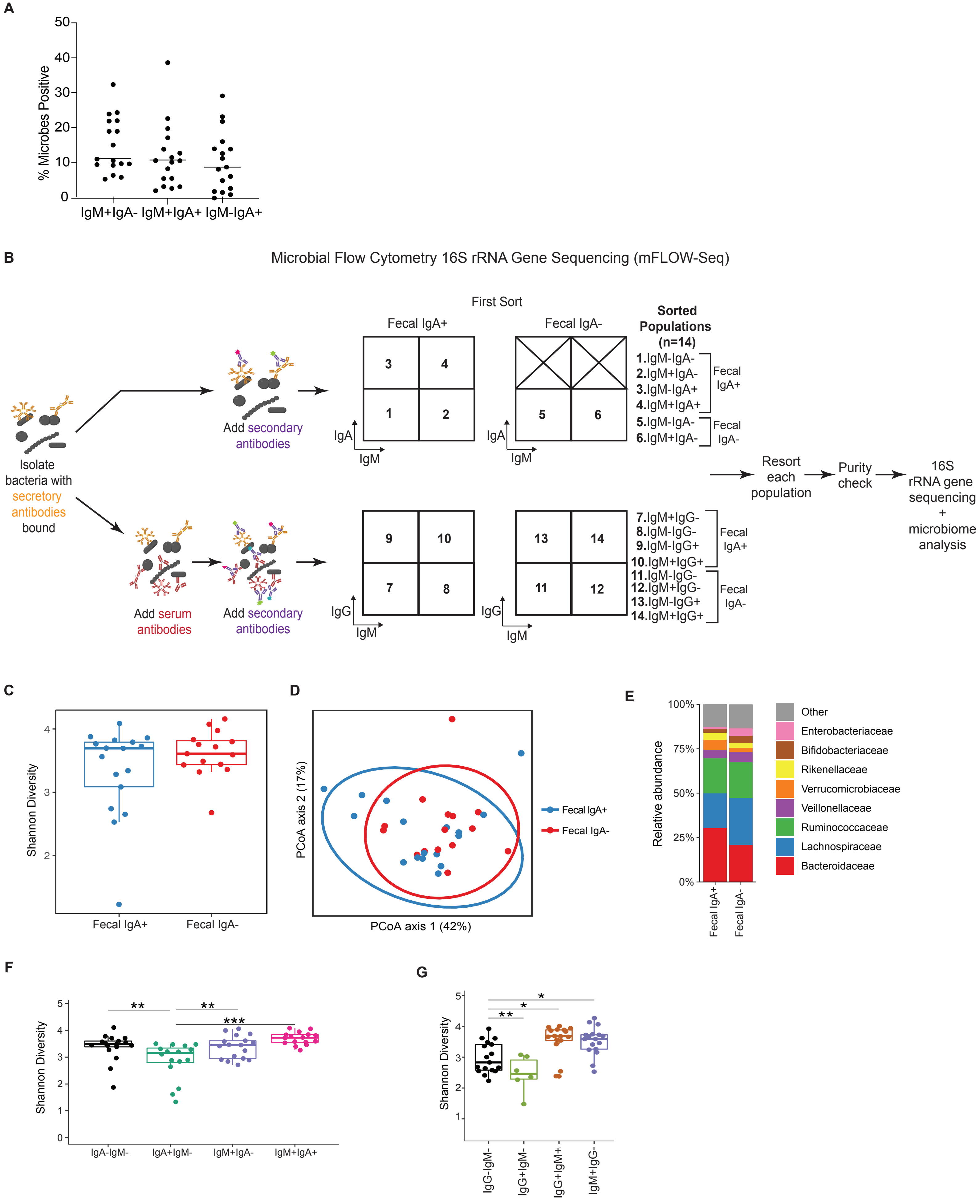

**Figure S2. Secretory IgA and IgM target an overlapping set of microbes in healthy children, related to Figure 1.**

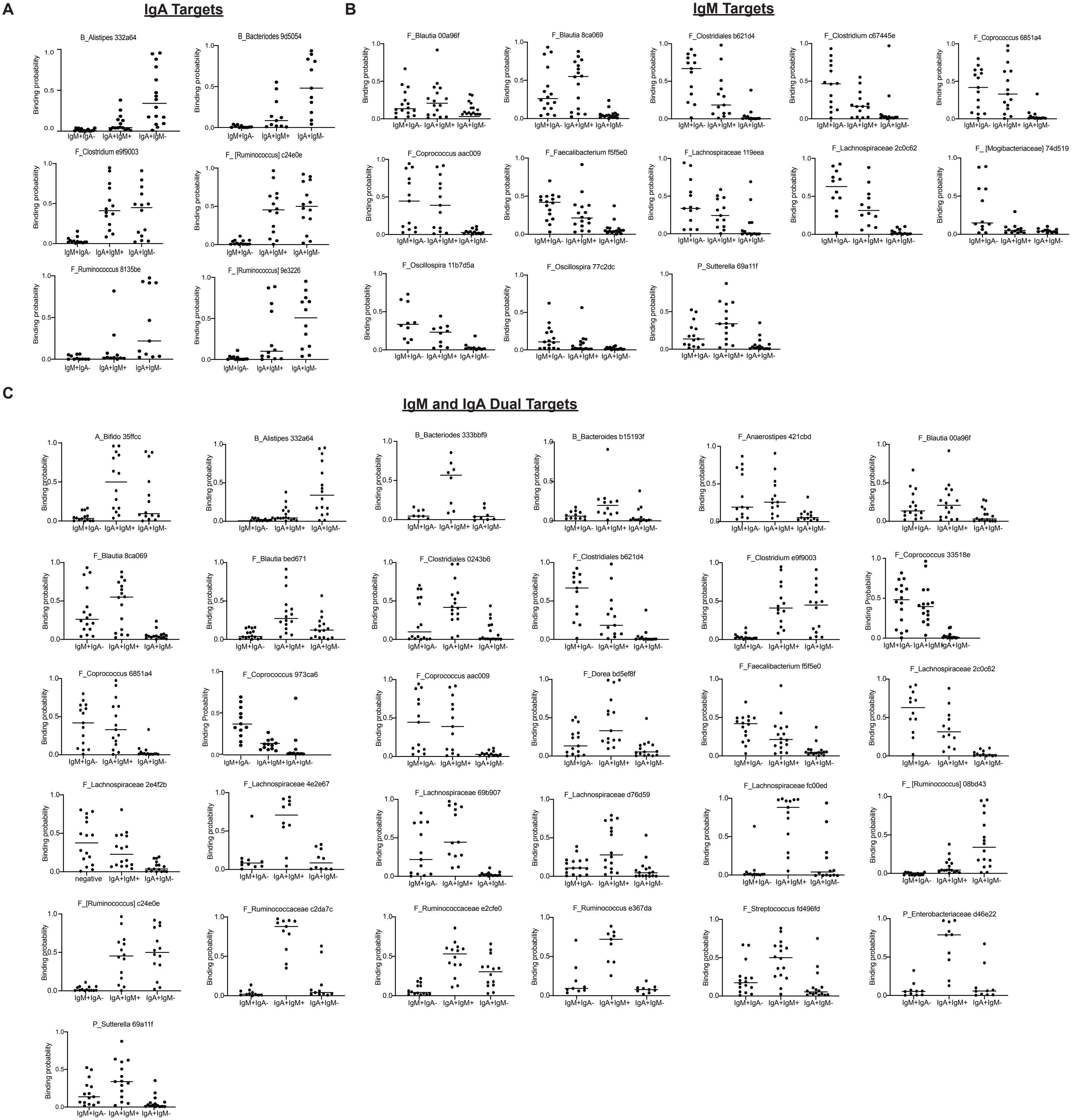

**D**

**IgM Targets**

**IgM+IgA- > IgM-IgA+**
**N=15**

F_Clostridium c67445e
F_Lachnospiraceae 119eea
F_Oscillospira 11b7d5a
F_Oscillospira 77c2dc
F_ [Mogibacteriaceae] 74d519

**IgM+IgA- >IgM-IgA+ AND IgM+IgA+ >IgM-IgA+**
**N=10**

F_Blautia 00a96f
F_Blautia 8ca0b9
F_Clostridiales b621d4
F_Coprococcus 33518e
F_Coprococcus 6851a4
F_Coprococcus 973ca6
F_Coprococcus aac009
F_Faecalibacterium f5f5e0
F_Lachnospiraceae 2c0c62
P_Sutterella 69a11f

**IgM+IgA+ >IgM-IgA+**
**N=25**

B_Bacteroides 333bbf9
B_Bacteroides b15193f
F_Anaerostipes 421cbd
F_Clostridiales 0243b6
F_Dorea bd5ef8f
F_Lachnospiraceae 2e4f2b
F_Lachnospiraceae 4e2e67
F_Lachnospiraceae 69b907
F_Lachnospiraceae d76d59
F_Lachnospiraceae fc00ed
F_Peptostreptococcaceae e335f74
F_Ruminococcaceae c2da7c
F_Ruminococcus e367da
F_Streptococcus fd496fd
P_Enterobacteriaceae d46e22

FDR<0.05

**IgA Targets**

**IgM-IgA+ > IgA-IgM+**
**N=8**

B_Bacteroides 9d5054
F_ [Ruminococcus] 9e3226
F_Ruminococcus 8135be

**IgM-IgA+ >IgA-IgM+ AND IgM+IgA+ >IgA-IgM+**
**N=5**

B_Allstipes 332a64
F_[Ruminococcus] 08bd43
F_Ruminococcus c24e0e
F_Clostridium e9f9003
F_Ruminococcaceae e2cfe0

**IgM+IgA+ >IgA-IgM+**
**N=12**

A_Bifidobacterium 35ffcc
F_Blautia bed671
F_Lachnospiraceae fc00ed
F_Peptostreptococcaceae e335f7
F_Ruminococcaceae c2da7c
P_Enterobacteriaceae d46e22
V_Akkermansia 7b28c2

FDR<0.05

**Figure S3. Systemic IgM and IgG target an overlapping set of microbes in healthy children, related to Figure 1**

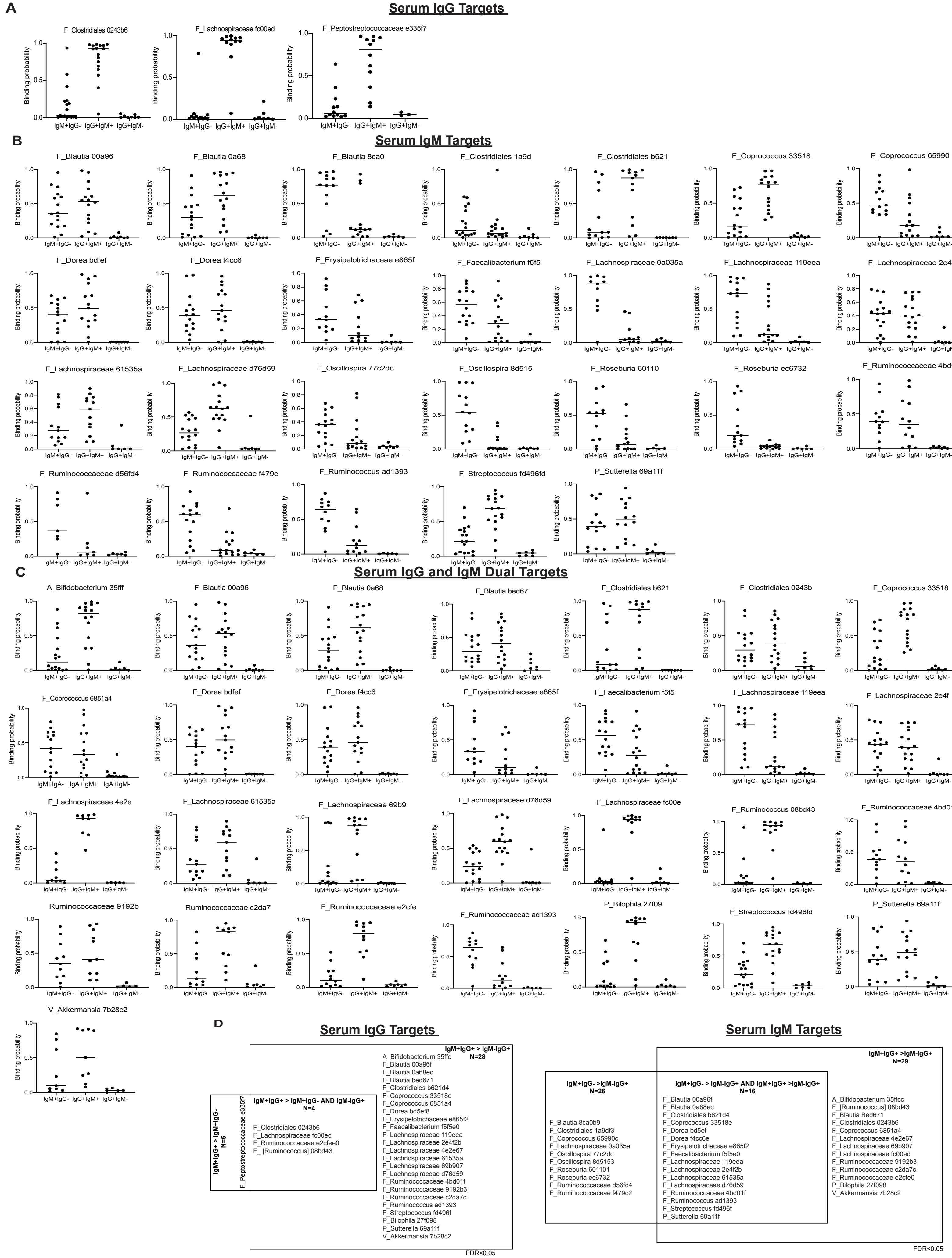

**Figure S4. Serum IgA concentration does not correlate to fecal IgA concentration or the percent of bacteria bound by IgA, related to Figure 3.**

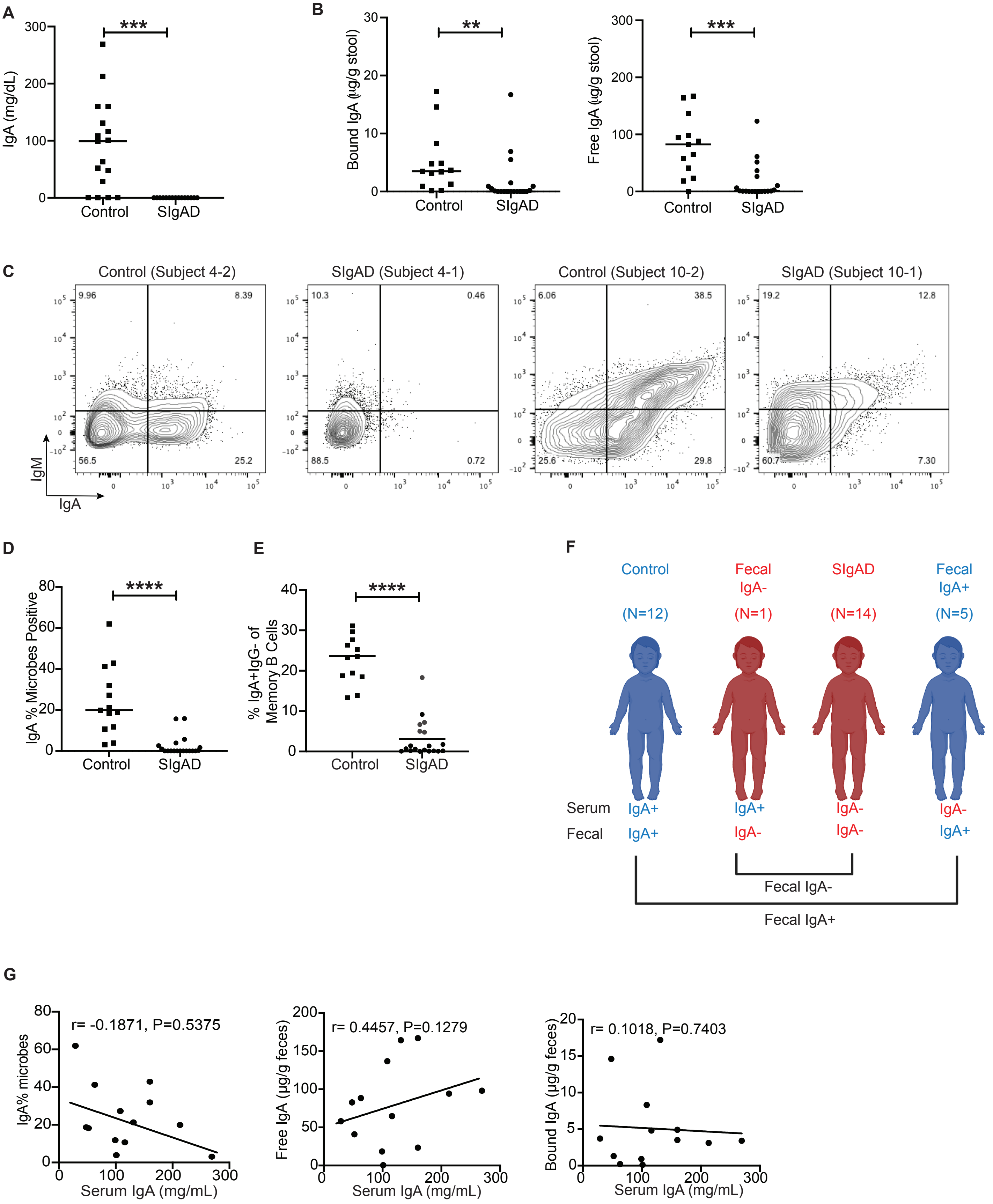

**Figure S5. Serum IgG binds to a higher proportion of microbes in fecal IgA deficient humans and mice compared to healthies, related to Figure 4.**

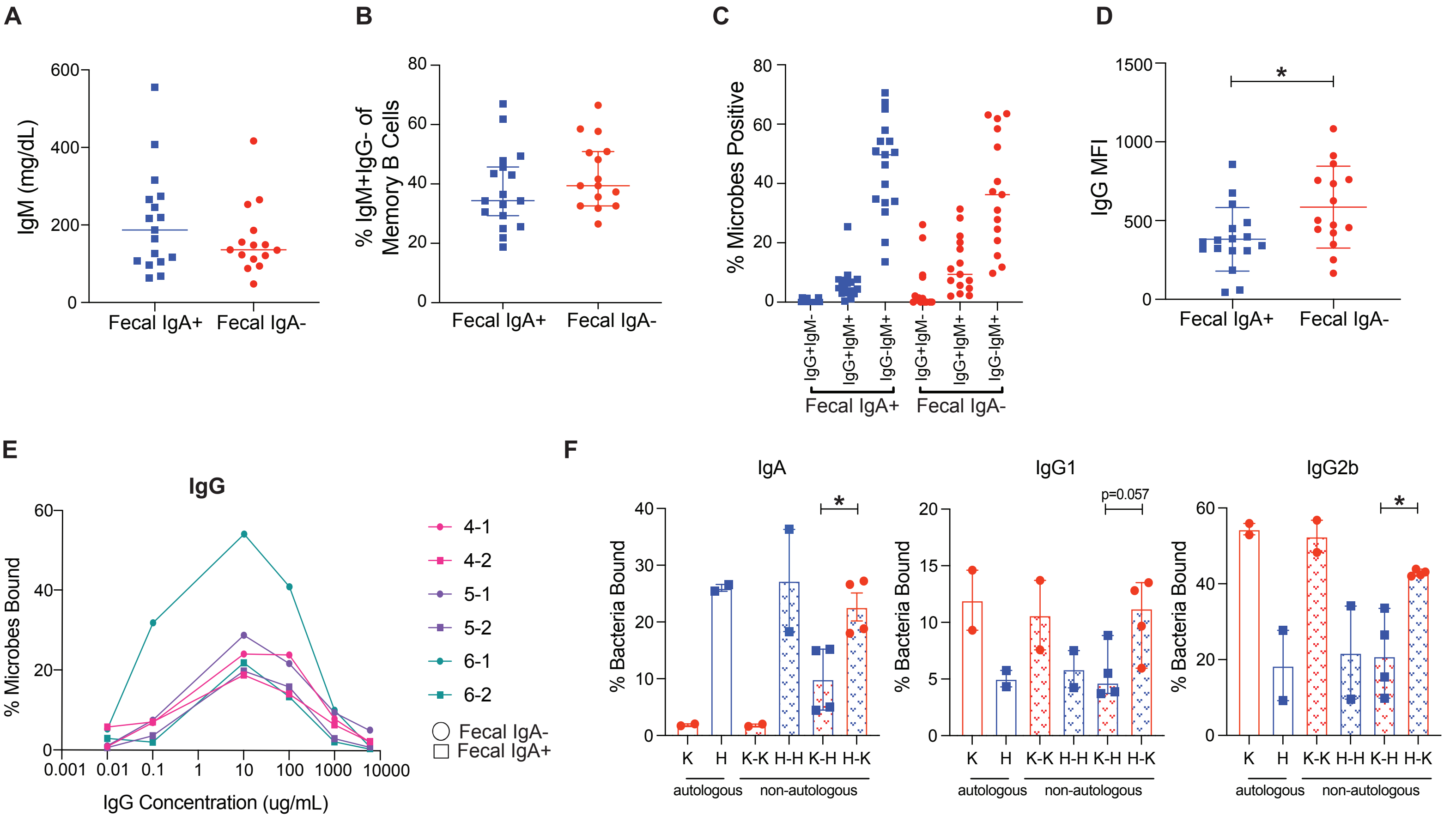

**Figure S6. Cellular immune-phenotyping in IgA deficient humans and mice, related to Figure 6.**

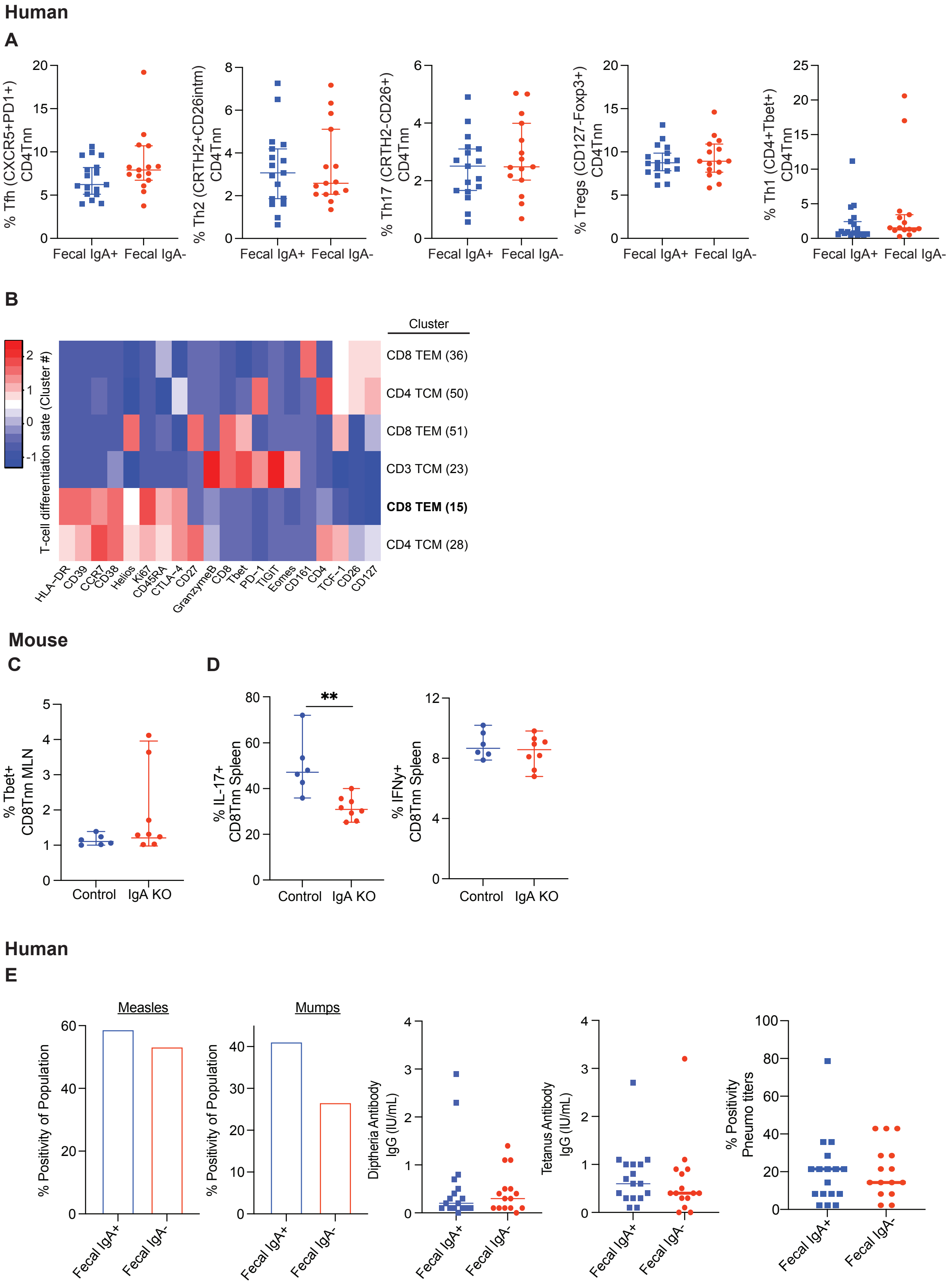

**Figure S7. Fecal IgA deficient humans and mice display serum cytokine and chemokine dysregulation, related to Figure 7.**

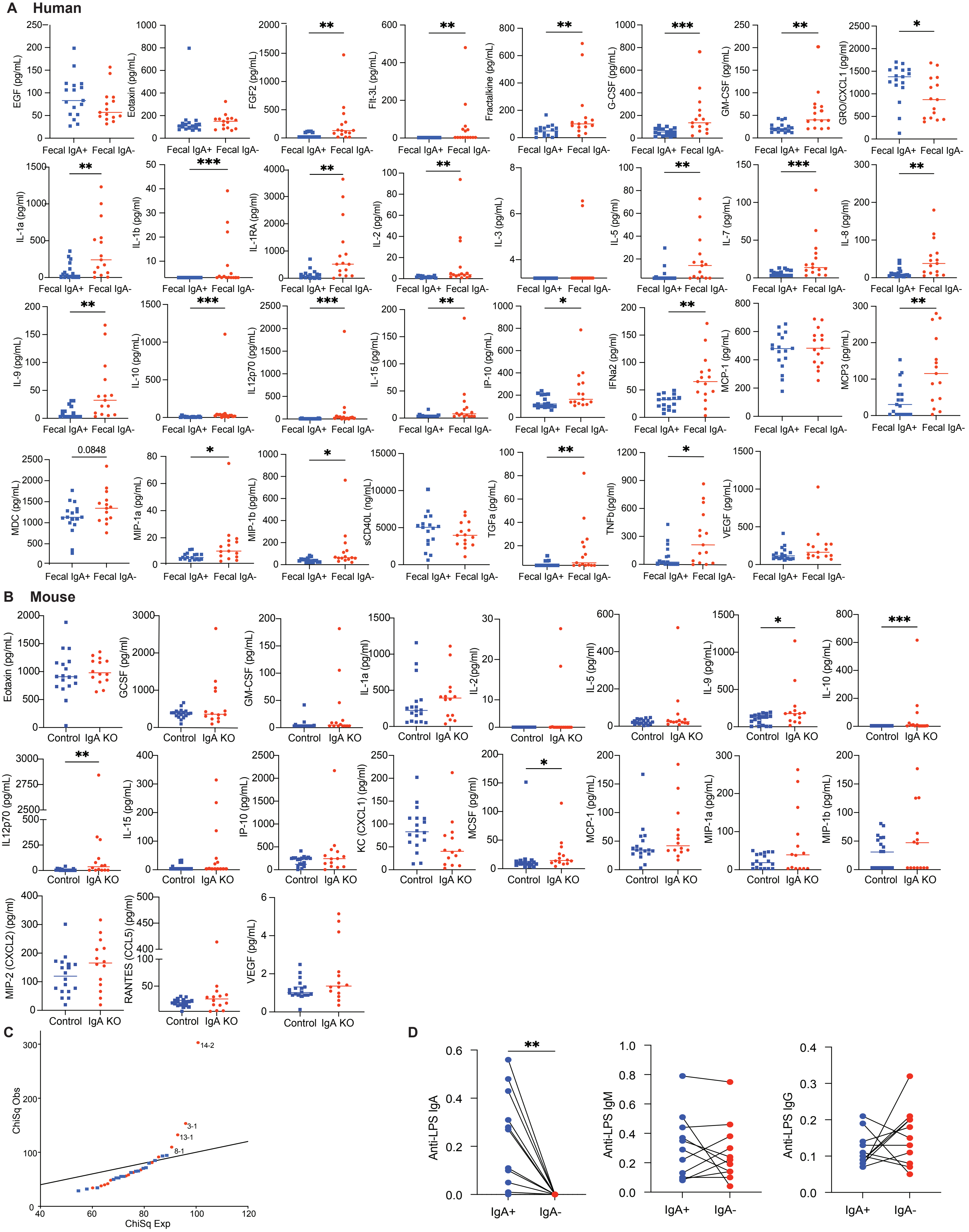

**Table S1. Demographics**

| Characteristic | Overall N=32 | Selective IgA deficiency N=15 | Control N=17 |
|---|---|---|---|
| ***Demographics*** | | | |
| **Age, years** | | | |
| Median(IQR) | 13(8-19) | 13.5(10.3-17) | 13(9.8-17) |
| **Sex** | | | |
| Male, n(%) | 13(40.6%) | 5(33.3%) | 8(47.1%) |
| Female, n(%) | 19(59.4%) | 10(66.7%) | 9(52.9%) |
| **BMI** | | | |
| **Percentile*** | | | |
| Median(IQR) | 70.5(20-91) | 70.5(35.5-80.8) | 70.5(38.5-81) |

*Percentile is based on the Center for Disease Control's weight percentile calculator for children 2-21 years old. Any enrolled subject > 21 years of age did not have a percentile calculated and were not considered in the median and interquartile range.
*Two subjects from the same family did not have vitals collected at their study visit.
IQR, Interquartile range

**Table S2. Beta Diversity of Sorted Populations**

| IgM and IgA Comparison (Group) | Comparison of Sorted Populations | Df | F Model | $R^2$ | p value |
|---|---|---|---|---|---|
| Fecal IgA+ | IgM+IgA- v. IgM- IgA- | 1 | 6 | 0.16 | 0.001 |
| Fecal IgA+ | IgM-IgA+ v. IgM-IgA- | 1 | 1.1 | 0.0036 | 0.045 |
| Fecal IgA+ | IgM+IgA+ v. IgM-IgA- | 1 | 3.3 | 0.095 | 0.003 |
| Fecal IgA+ | IgM+IgA- v. IgM+IgA+ | 1 | 4.2 | 0.12 | 0.001 |
| Fecal IgA+ | IgM-IgA+ v. IgM+IgA+ | 1 | 1.5 | 0.047 | 0.018 |
| Fecal IgA+ | IgM-IgA+ v. IgM+IgA- | 1 | 3.5 | 0.1 | 0.002 |

| IgM and IgG Comparison (Group) | Comparison of Sorted Populations | Df | F Model | $R^2$ | p value |
|---|---|---|---|---|---|
| Fecal IgA+ | IgM+IgG- v. IgM-IgG- | 1 | 3.9 | 0.11 | 0.001 |
| Fecal IgA+ | IgM-IgG+ v. IgM-IgG- | 1 | 2.7 | 0.11 | 0.045 |
| Fecal IgA+ | IgM+IgG+ v. IgM-IgG- | 1 | 4.7 | 0.13 | 0.003 |
| Fecal IgA+ | IgM+IgG- v. IgM+IgG+ | 1 | 1.7 | 0.073 | 0.001 |
| Fecal IgA+ | IgM+IgG- v. IgM+IgG+ | 1 | 2.1 | 0.063 | 0.018 |
| Fecal IgA+ | IgM-IgG+ v. IgM+IgG+ | 1 | 2.8 | 0.11 | 0.002 |

**Table S3. Antibody Binding Probabilities for IgM, IgA and IgG**

| Name | ASV | IgM binding probability | IgA binding probability | IgG binding probability | Top IgG targets |
|---|---|---|---|---|---|
| Firmicutes [Ruminococcus] | c24e0e | 0.47 | 0.88 | 0.54 | |
| ***Firmicutes [Ruminococcus]*** | ***08bd43*** | ***0.74*** | ***0.85*** | ***0.73*** | ***Y*** |
| ***Firmicutes Lachnospiraceae*** | ***fc00ed*** | ***0.73*** | ***0.83*** | ***0.73*** | ***Y*** |
| Firmicutes Clostridium | e9f900 | 0.50 | 0.80 | 0.64 | |
| Firmicutes Ruminococcaceae | c2da7c | 0.72 | 0.79 | 0.55 | |
| Actinobacteria Bifidobacterium | 35ffcc | 0.54 | 0.75 | 0.68 | |
| ***Firmicutes Ruminococcaceae*** | ***e2cfe0*** | ***0.55*** | ***0.74*** | ***0.62*** | ***Y*** |
| ***Firmicutes Peptostreptococcaceae*** | ***e335f7*** | ***0.72*** | ***0.70*** | ***0.63*** | ***Y*** |
| Firmicutes [Ruminococcus] | 9e3226 | 0.36 | 0.70 | 0.42 | |
| Proteobacteria Enterobacteriaceae | d46e22 | 0.65 | 0.69 | 0.51 | |
| Firmicutes Lachnospiraceae | 4e2e67 | 0.67 | 0.66 | 0.66 | |
| Actinobacteria Bifidobacterium | 315ca0 | 0.65 | 0.65 | 0.47 | |
| Firmicutes Lachnospiraceae | fcffed | 0.52 | 0.65 | 0.49 | |
| Verrucomicrobia Akkermansia | 7b28c2 | 0.52 | 0.64 | 0.47 | |
| Firmicutes Ruminococcus | e367da | 0.69 | 0.63 | 0.53 | |
| Bacteroidetes Rikenellaceae | 506efa | 0.40 | 0.62 | 0.34 | |
| Bacteroidetes Bacteroides | 9d5054 | 0.28 | 0.62 | 0.47 | |
| Firmicutes Dorea | 7af6fc | 0.59 | 0.61 | 0.56 | |
| Firmicutes Clostridium | 9acb55 | 0.47 | 0.61 | 0.51 | |
| Firmicutes Streptococcus | fd496f | 0.70 | 0.60 | 0.65 | |
| Firmicutes Faecalibacterium | c48070 | 0.46 | 0.58 | 0.45 | |
| Firmicutes Lachnospiraceae | 69b907 | 0.82 | 0.58 | 0.60 | |
| Firmicutes Turicibacter | 26c6e3 | 0.59 | 0.57 | 0.42 | |
| Firmicutes Ruminococcaceae | 3f49f2 | 0.56 | 0.57 | 0.52 | |
| ***Firmicutes Clostridiales*** | ***0243b6*** | ***0.72*** | ***0.57*** | ***0.76*** | ***Y*** |
| Firmicutes Oscillospira | 32cc17 | 0.56 | 0.56 | 0.52 | |
| Firmicutes Blautia | c6c3ab | 0.40 | 0.56 | 0.65 | |
| Firmicutes Dorea | bd5ef8 | 0.64 | 0.55 | 0.50 | |
| Firmicutes Peptostreptococcaceae | 642067 | 0.53 | 0.55 | 0.46 | |
| Bacteroidetes Alistipes | 8795c5 | 0.39 | 0.55 | 0.42 | |
| Firmicutes Oscillospira | 68d79c | 0.61 | 0.53 | 0.51 | |
| Firmicutes Ruminococcus | 8135be | 0.25 | 0.53 | 0.37 | |
| Firmicutes Erysipelotrichaceae | e865f2 | 0.38 | 0.52 | 0.28 | |
| Bacteroidetes Bacteroides | 333bbf | 0.52 | 0.52 | 0.43 | |
| Bacteroidetes Bacteroides | 2c9829 | 0.41 | 0.52 | 0.53 | |
| Firmicutes Blautia | 8ca0b9 | 0.82 | 0.51 | 0.29 | |
| Actinobacteria Collinsella | 35cbb3 | 0.49 | 0.50 | 0.44 | |
| Bacteroidetes Bacteroides | d900eb | 0.46 | 0.50 | 0.42 | |
| Firmicutes Ruminococcaceae | 410e1e | 0.46 | 0.50 | 0.38 | |

| | | | | |
|---|---|---|---|---|
| Bacteroidetes Alistipes | 332a64 | 0.11 | 0.50 | 0.32 |
| Firmicutes Clostridiales | f0015c | 0.44 | 0.50 | 0.45 |
| Firmicutes Blautia | bed671 | 0.41 | 0.49 | 0.46 |
| Firmicutes Faecalibacterium | 8e175a | 0.27 | 0.49 | 0.39 |
| Firmicutes Ruminococcaceae | 4bd01f | 0.53 | 0.48 | 0.38 |
| Firmicutes Blautia | 0a68ec | 0.58 | 0.47 | 0.58 |
| Bacteroidetes Bacteroides | fd44d4 | 0.40 | 0.46 | 0.39 |
| Firmicutes Lachnospiraceae | d76d59 | 0.52 | 0.46 | 0.60 |
| Firmicutes Dorea | f4cc6e | 0.57 | 0.46 | 0.48 |
| Bacteroidetes Odoribacter | a730ef | 0.43 | 0.46 | 0.47 |
| Firmicutes Coprococcus | aac009 | 0.78 | 0.46 | 0.54 |
| Firmicutes Coprococcus | 33518e | 0.86 | 0.45 | 0.65 |
| Firmicutes Coprococcus | 6851a4 | 0.78 | 0.45 | 0.70 |
| Bacteroidetes Alistipes | 742e1f | 0.29 | 0.44 | 0.51 |
| Firmicutes Anaerostipes | 421cbd | 0.67 | 0.44 | 0.37 |
| Proteobacteria Sutterella | 69a11f | 0.55 | 0.43 | 0.49 |
| Firmicutes Oscillospira | 3219af | 0.42 | 0.43 | 0.50 |
| Firmicutes Lachnospiraceae | 61535a | 0.49 | 0.43 | 0.51 |
| Firmicutes Lachnospiraceae | 0a035a | 0.48 | 0.42 | 0.22 |
| Bacteroidetes Bacteroides | 824b9c | 0.42 | 0.42 | 0.37 |
| Bacteroidetes Rikenellaceae | 51d4f1 | 0.43 | 0.42 | 0.34 |
| Firmicutes Coprococcus | 65990c | 0.46 | 0.42 | 0.31 |
| Firmicutes Lachnospiraceae | 2c0c62 | 0.83 | 0.42 | 0.52 |
| Firmicutes Lachnospiraceae | caea32 | 0.42 | 0.41 | 0.45 |
| Firmicutes Lachnospiraceae | 119eea | 0.65 | 0.41 | 0.33 |
| Bacteroidetes Parabacteroides | d8e7f9 | 0.33 | 0.41 | 0.35 |
| Bacteroidetes Bacteroides | 1b8dec | 0.40 | 0.41 | 0.37 |
| Firmicutes Ruminococcaceae | d56fd4 | 0.47 | 0.41 | 0.29 |
| Bacteroidetes Bacteroides | 836cd5 | 0.27 | 0.40 | 0.36 |
| Firmicutes Ruminococcus | 149465 | 0.52 | 0.40 | 0.33 |
| Firmicutes Lachnospiraceae | b0e56c | 0.42 | 0.39 | 0.31 |
| Firmicutes Ruminococcaceae | 9192b3 | 0.54 | 0.39 | 0.47 |
| Firmicutes Roseburia | ab4040 | 0.44 | 0.38 | 0.35 |
| Firmicutes Dialister | 818434 | 0.46 | 0.38 | 0.66 |
| Bacteroidetes Bacteroides | bf6114 | 0.33 | 0.38 | 0.27 |
| Firmicutes Clostridiales | b621d4 | 0.80 | 0.37 | 0.59 |
| Firmicutes Christensenellaceae | a72bc4 | 0.44 | 0.37 | 0.49 |
| Proteobacteria Bilophila | 27f098 | 0.46 | 0.35 | 0.62 |
| Bacteroidetes Bacteroides | b15193 | 0.35 | 0.35 | 0.39 |
| Bacteroidetes Bacteroides | 76b39e | 0.28 | 0.35 | 0.33 |
| Firmicutes Lachnospiraceae | d622c6 | 0.31 | 0.35 | 0.43 |
| Firmicutes Lachnospiraceae | 662a40 | 0.51 | 0.34 | 0.41 |
| Firmicutes Lachnospiraceae | 2e4f2b | 0.55 | 0.34 | 0.43 |
| Firmicutes Faecalibacterium | f5f5e0 | 0.64 | 0.34 | 0.33 |

| | | | | |
|---|---|---|---|---|
| Firmicutes Faecalibacterium | 597771 | 0.28 | 0.34 | 0.26 |
| Firmicutes Oscillospira | 11b7d5 | 0.54 | 0.34 | 0.44 |
| Firmicutes Oscillospira | 5d08d1 | 0.39 | 0.33 | 0.42 |
| Firmicutes Clostridium | c67445 | 0.62 | 0.33 | 0.35 |
| Firmicutes Blautia | 00a96f | 0.45 | 0.33 | 0.48 |
| Firmicutes Phascolarctobacterium | 1569f7 | 0.42 | 0.32 | 0.32 |
| Firmicutes Ruminococcus | 596db8 | 0.40 | 0.32 | 0.44 |
| Firmicutes Ruminococcaceae | f479c2 | 0.42 | 0.31 | 0.21 |
| Firmicutes Ruminococcus | d9eea8 | 0.25 | 0.31 | 0.39 |
| Bacteroidetes Bacteroides | 99deb3 | 0.17 | 0.28 | 0.23 |
| Firmicutes Roseburia | 601101 | 0.46 | 0.28 | 0.22 |
| Firmicutes Butyricicoccus | a18c0c | 0.46 | 0.28 | 0.25 |
| Firmicutes Faecalibacterium | 86ff0e | 0.33 | 0.28 | 0.25 |
| Firmicutes Coprococcus | 973ca6 | 0.50 | 0.27 | 0.42 |
| Firmicutes Gemmiger | 9639a3 | 0.22 | 0.24 | 0.36 |
| Firmicutes Clostridiales | 1a9df3 | 0.17 | 0.21 | 0.17 |
| Firmicutes [Mogibacteriaceae] | 74d519 | 0.43 | 0.21 | 0.23 |
| Firmicutes Ruminococcus | ad1393 | 0.42 | 0.21 | 0.25 |
| Firmicutes Roseburia | 263e41 | 0.14 | 0.18 | 0.11 |
| Firmicutes Oscillospira | f98dab | 0.20 | 0.18 | 0.21 |
| Firmicutes Oscillospira | 5fbc48 | 0.17 | 0.18 | 0.17 |
| Firmicutes Oscillospira | 8d5153 | 0.33 | 0.17 | 0.16 |
| Firmicutes Roseburia | ec6732 | 0.23 | 0.16 | 0.12 |
| Firmicutes Oscillospira | 77c2dc | 0.26 | 0.13 | 0.23 |

**Bold italicized ASVs indicate the top 5 IgG targeted microbes.**

**Table S4. Defining IgA status based on serum IgA concentration, fecal IgA concentration and % IgA binding to commensal microbes**

| Subject | Family | Serum IgA (mg/dL) | Serum IgG (mg/dL) | % of IgA+ B cells in blood | Clinical Diagnosis (based on serum IgA) | Free Fecal IgA (ug/g) | Bound Fecal IgA (ug/g) | % of Fecal Microbes bound by IgA | Fecal IgA status* | Comment |
|---|---|---|---|---|---|---|---|---|---|---|
| **1-1** | **1** | **<7** | 1410 | **7.2** | **Selective IgA def** | **123** | **17** | **15.8** | **IgA+** | **Serum IgA-, Stool IgA+** |
| 1-2 | 1 | <7 | 1280 | 9.2 | Selective IgA def | 3 | 1 | 2.6 | IgA def | |
| **2-1** | **2** | **<26** | **1020** | **4.8** | **Selective IgA def** | **61** | **7** | **1.7** | **IgA+** | **Serum IgA-, Stool IgA+** |
| **2-2** | **2** | **<7** | **920** | **1.4** | **Selective IgA def** | **37** | **6** | **3.9** | **IgA+** | **Serum IgA-, Stool IgA+** |
| 3-1 | 3 | <7 | 1600 | 0.4 | Selective IgA def | 10 | 1 | 1.1 | IgA def | |
| 4-1 | 4 | <7 | 1250 | 0.1 | Selective IgA def | 1 | 0 | 0.0 | IgA def | |
| 5-1 | 5 | <7 | 1490 | 0.2 | Selective IgA def | 6.2 | 0 | 0.0 | IgA def | |
| 6-1 | 6 | <7 | 1100 | 0.6 | Selective IgA def | 0 | 0 | 0.0 | IgA def | |
| 7-1 | 7 | <7 | 1220 | 1.7 | Selective IgA def | 0 | 0 | 0.0 | IgA def | |
| 8-1 | 8 | <7 | 1690 | 5.0 | Selective IgA def | 0 | 0 | 0.0 | IgA def | |
| 8-2 | **8** | **<7** | **1120** | **1.4** | **Selective IgA def** | **52** | **2** | **5.7** | **IgA+** | **Serum IgA-, Stool IgA+** |
| 9-1 | 9 | <7 | 1470 | 0.0 | Selective IgA def | 1 | 0 | 0.3 | IgA def | |
| 10-1 | **10** | **<6** | **1290** | **0.2** | **Selective IgA def** | **26** | **1** | **15.7** | **IgA+** | **Serum IgA-, Stool IgA+** |
| 11-1 | 11 | <7 | 1020 | 0.1 | Selective IgA def | 1 | 0 | 0.0 | IgA def | |
| 12-1 | 12 | <6 | 682 | 18.3 | Selective IgA def | 1 | 0 | 0.0 | IgA def | |
| 13-1 | 13 | <6 | 1230 | 0.7 | Selective IgA def | 1 | 0 | 0.0 | IgA def | |
| 14-1 | 14 | <7 | 1670 | 0.1 | Selective IgA def | 1 | 0 | 0.0 | IgA def | |
| 14-2 | 14 | <7 | 1410 | 0.1 | Selective IgA def | 0 | 0 | 0.0 | IgA def | |

| | | | | | | | | | | |
|---|---|---|---|---|---|---|---|---|---|---|
| 15-1 | 15 | <6 | 1500 | 6.7 | Selective IgA def | 0 | 0 | 0.0 | IgA def | |
| **3-2** | **3** | **101** | **907** | **32.7** | **Control** | **0** | **0** | **3.9** | **IgA def** | **Serum IgA+, Stool IgA-** |
| 4-2 | 4 | 48 | 817 | 29.6 | Control | 83 | 15 | 31.9 | IgA+ | |
| 5-2 | 5 | 160 | 982 | 23.3 | Control | 167 | 4 | 18.7 | IgA+ | |
| 6-2 | 6 | 108 | 743 | 18.7 | Control | 137 | 8 | 42.9 | IgA+ | |
| 7-2 | 7 | 29 | 886 | 13.3 | Control | 58 | 4 | 3.1 | IgA+ | |
| 9-2 | 9 | 99 | 749 | 26.3 | Control | 18 | 1 | 27.3 | IgA+ | |
| 9-3 | 9 | 52 | 855 | 13.9 | Control | 41 | 1 | 19.9 | IgA+ | |
| 10-2 | 10 | 131 | 1220 | 31.1 | Control | 164 | 17 | 61.9 | IgA+ | |
| 11-2 | 11 | 116 | 839 | 19.4 | Control | 65 | 5 | 11.8 | IgA+ | |
| 12-2 | 12 | 160 | 1200 | 25.3 | Control | 23 | 5 | 18.2 | IgA+ | |
| 12-3 | 12 | 269 | 1730 | 23.6 | Control | 98 | 3 | 21.3 | IgA+ | |
| 13-2 | 13 | 213 | 1300 | 27.6 | Control | 94 | 3 | 10.7 | IgA+ | |
| 15-2 | 15 | 63 | 996 | 18.5 | Control | 88 | 0 | 41.2 | IgA+ | |

***Fecal IgA deficiency (IgA def)** requires <u>all</u> of the following 3 conditions to be met:

1. Less than or equal to 1 ug/g IgA bound to bacteria
2. Less than 10 ug/g free fecal IgA
3. Less than 5% IgA bound microbes on mFLOW.

**Fecal IgA sufficient (IgA+)** requires <u>at least one</u> of the following 3 conditions be met:

1. Greater than 1 ug/g IgA bound to bacteria
2. Greater than 10 ug/g free fecal IgA
3. Greater than 5% IgA bound microbes on mFLOW.

Bolded individuals indicate discordance between serum and fecal IgA status.
